# Declining HIV incidence in sub-Saharan Africa: a systematic review and meta-analysis of empiric data

**DOI:** 10.1101/2020.12.08.20246066

**Authors:** Keya Joshi, Justin Lessler, Oluwasolape Olawore, Gideon Loevinsohn, Sophrena Bushey, Aaron A.R. Tobian, M. Kate Grabowski

## Abstract

**Background:** UNAIDS models suggest HIV incidence is declining in sub-Saharan Africa; however, it is unclear whether modeled trends are supported by empirical evidence.

**Methods:** We conducted a systematic review and meta-analysis of adult HIV incidence data from sub-Saharan Africa by searching Embase, Scopus, PubMed, and OVID databases and technical reports published between January 1, 2010 and July 23, 2019. We included studies that directly measured incidence from blood samples. Incidence data were abstracted according to population risk group, geographic location, sex, intervention arm, and calendar period. Weighted regression models were used to assess incidence trends across general population studies by sex. We also identified studies reporting ≥3 incidence measurements since 2010 and assessed trends within them.

**Findings:** 292 studies met inclusion criteria. Most studies were conducted in South Africa (n=102), Uganda (n=46), and Kenya (n=41); there were 27 countries with no published incidence data, most in western and central Africa. Across general population studies, average annual incidence declines since 2010 were 0.16/1000 person-years (95%CI:0.06-0.26;p=0.004) among men and 0.16/1000 person-years (95%CI: −0.01-0.33;p=0.060) among women in eastern Africa, and 0.25/1000 person-years (95%CI:0.17-034;p<0.0001) among men and 0.42/1000 person-years (95%CI:0.23-0.62;p=0.0002) among women in southern Africa. In 9/10 studies with multiple measurements, incidence declined over time. Incidence was typically higher in women than men (median ratio=1.45, IQR: 1.12-1.83) with increasing sex disparity over time.

**Interpretation:** Empirical incidence data show the rate of new HIV infections is declining in eastern and southern Africa. However, recent incidence data are non-existent or very limited for many countries, particularly in western and central Africa.

**Funding:** National Institute of Allergy and Infectious Diseases

## INTRODUCTION

Nearly 40 years into the HIV pandemic, sub-Saharan Africa remains the global epicenter of HIV transmission, accounting for 57% of all new infections in 2019^1^. Within most African countries, transmission continues to be generalized with substantial rates of new infections outside of clearly defined risk groups, particularly in eastern and southern Africa^1^. Widespread community transmission within the region has spurred substantial investment in population-based infection control measures, including biomedical and behavioral interventions. Domestic and international donors, including the Global Fund to Fight AIDS, Tuberculosis, and Malaria and the United States President’s Emergency Plan for AIDS Relief (PEPFAR), continue to support HIV programming in sub-Saharan African with significant financial and human resources investment in large-scale HIV testing, prevention, and treatment programs. However, global funding for HIV programming within Africa is waning, potentially jeopardizing progress towards UNAIDS fast track goals to end the African epidemic by 2030^2^. Measuring the impact of programmatic investments to control HIV is critical for demonstrating headway, appropriately targeting future funding and other resources, and ensuring a sustained global commitment to ending the HIV pandemic.

Key epidemiologic metrics of HIV control include absolute rates of HIV incidence and AIDS-related deaths and percent reductions in new infections and AIDS-related deaths among other benchmarks^3,4^. While each of these control criteria provide key insights into epidemic trajectories, HIV incidence is arguably the most important of these metrics because it is less impacted by fluctuations in non-HIV mortality and more comparable across different population and geographic settings^4^. However, directly observed HIV incidence measurements are difficult to obtain partly due to delays in testing following infection, limited availability of testing to detect early HIV infections, and difficulty enumerating and following populations at risk. Thus, data on HIV incidence trends in sub-Saharan Africa predominantly come from UNAIDS mathematical models which use demographic, HIV prevalence, and programmatic data^5–8^. Based on these models, UNAIDS reports HIV incidence is steadily declining across sub-Saharan Africa. However, there are limited direct empirical data supporting these modeled downward trends. The last comprehensive systematic review of the empiric HIV incidence literature from sub-Saharan Africa was published in 2009^9^. This prior review, including only 57 studies, found that HIV incidence data from western and central Africa were rare and that estimates varied substantially within country and across population risk groups: no trend data were analyzed.

Here, we provide an update on empirical HIV incidence data published over the last decade. Our systematic review and meta-analysis includes all studies reporting directly observed HIV incidence measurements obtained using serologic assays either through prospective or cross-sectional assessments. Data were stratified by gender and risk group where possible. Regional variation in incidence data are reported along with trends over time across and within studies, by sex, and across various population risk groups, including men who have sex with men (MSM) and sex workers.

## METHODS

### Screening and Extraction Protocol

We performed a systematic literature review and meta-analysis to identify studies on HIV incidence from sub-Saharan Africa published between January 1, 2010 and July 23, 2019. We searched PubMed, Embase, Scopus, and OVID global health databases for peer-reviewed articles reporting directly observed (i.e. empirical) estimates of HIV incidence measured through either prospective repeat testing or cross-sectional HIV incidence testing of blood samples. We used the following search terms “HIV”, “incidence”, and “Africa” as medical subject heading (MESH) terms. Additional clinical synonyms and alternative spellings were also included in the search (see supplemental methods for full search criteria). Technical reports measuring population-level HIV incidence, but not published in peer-reviewed journals, such as the population-based HIV impact assessment (PHIA) surveys, were also included.

Our analysis included peer-reviewed studies and technical reports including national or subnational empiric estimates of HIV incidence from sub-Saharan Africa. We excluded studies of HIV not in humans, studies reporting incidence estimates for children < 15 years of age only, studies not in English, perspectives, opinions, commentaries, case-control studies, studies measuring impact of post-exposure prophylaxis among health care workers (HCW), cross-sectional incidence studies only including HIV-positive persons, and incidence estimates based on mathematical models. We further excluded studies with incidence estimates based on less than 50 total participants or less than 50 person-years at risk

After removal of duplicates, all studies identified through the database search were uploaded to the Covidence systematic review management software and then screened by two independent reviewers (KJ, SB, GL, OO)^10^. Studies first went through a title and abstract review. Those deemed eligible went through further full text review. As defined in the protocol, any disagreement at each stage was resolved through consensus.

We extracted standardized data from each study including study design, study cohort, geographic location (ISO1-ISO3, where applicable), dates of data collection (i.e. the study period), incidence rate (including 95% confidence intervals [95%CI] and standard errors [SE]), cumulative incidence (including 95% CIs and SEs), person-years (pys) at risk, number of HIV seroconversions, number of individuals in the study overall and by sex and age where applicable, total numbers of HIV-negative and positive participants at baseline, and a description of the intervention if the study was a randomized clinical trial (RCT). Data from multi-center RCTs were disaggregated by site if possible. Incidence rates were further disaggregated by sex, age group/range, population risk group, and intervention arm when available. Population risk groups included sex workers (SW), men who have sex with men (MSM), transgender women (TGW), pregnant women (PW), serodiscordant couples (SDC), Lake Victoria fisherfolk (FF), other high risk populations (e.g., women with multiple sexual partners; bar workers), and general populations (i.e., geographically defined populations with no defining characteristics beyond location of residency). For studies measuring cross sectional incidence through either BED™ capture enzyme immunoassay (BED-CEIA) or Limiting Antigen avidity (LAg-Avidity) enzyme immunoassay, we also recorded information on the window period, false recency rate and optical density (OD) reported.

For incidence rates, all estimates were converted to number of seroconversions per 100 pys at risk. We did not assess study quality or perform a risk of bias assessment for three reasons: (1) rates of HIV incidence were expected to vary across geography and between risk groups, (2) our primary objective was to summarize incidence data as opposed to assessing the causal impact of any particular intervention or exposure, and (3) there was substantial heterogeneity in the methods used to obtain incidence estimates.

### Statistical Analysis

Studies were grouped into geographic regions as defined by the UN Statistics Division^11^. However, Malawi, Mozambique, Zambia and Zimbabwe were categorized as part of the southern region, and Sudan as part of the eastern region, due to the similarity of their epidemic, most notably subtype distribution, with other countries in those respective regions.

We summarized HIV incidence trends across general population studies over calendar time for eastern and southern Africa by using weighted loess regression for studies in which the calendar midpoint of the data collection period occurred on or after October 8, 2006 (i.e., the ≥25^th^ percentile of the distribution of data collection calendar midpoints). Weights corresponded to the study size (N) of the population. In order to estimate the rate of change in incidence since 2010, we fit weighted linear regression models weighted by study size for all studies in which the calendar midpoint of the data collection period was in 2010 or later. For countries where study size was not reported for a specific time-point, the most recent study size reported either at follow-up or baseline was used. All trend analyzes were stratified by sex. We excluded studies that only reported incidence estimates for men and women combined, age-specific incidence estimates for age bins less than 26 years, and studies that reported incidence estimates for non-general population subgroups. The 26 year cutoff was chosen because it was the smallest age bin including more than two thirds (>70%) of the adult population most at risk for HIV, typically defined as 15-49 years old. For studies where CIs were not reported, we derived exact confidence intervals based on a Poisson distribution. Studies that did not report an upper or lower age limit were included (e.g., studies that reported a minimum enrollment of 15 years but no max age enrollment), but these studies were excluded in sensitivity analyses. Point estimates were plotted at the midpoint of the data collection period (e.g. if a study was conducted from January 1, 2010 to December 31, 2010, the study midpoint would be July 2, 2010) with bars indicating the entire length of the collection period (e.g. January 1, 2010 and December 31, 2010). Trends in HIV incidence were not examined for western or central Africa or for other population risk groups given limited availability of incidence data.

We further summarized recent incidence trends over the last decade within study cohorts over time using linear regression models. This analysis was limited to studies that reported incidence estimates for at least three unique calendar periods since January 1, 2010. For studies that reported age-stratified estimates, we used incidence metrics for the widest age bin reported, but did not restrict based on age-bin size.

We constructed forest plots to summarize HIV incidence rates and 95% confidence intervals by region and risk group. For studies where incidence estimates were reported for multiple time periods, we only included the most recent estimate. Similarly, if multiple studies reported estimates from the same cohort, we used the most recent estimate available. For RCTs, we included the estimate from the control arm only. For studies reporting estimates disaggregated by type of hormonal contraceptive use, we included the incidence estimate among women using either no contraceptive, or non-hormonal contraceptives. For studies reporting both national and sub-national incidence data, estimates at the national level only were included. For general population studies, we only reported incidence estimates for age bins spanning 26 years or greater.

All analyses were conducted in R (version 3.6.2) and Stata (version 14).

## RESULTS

There were 34,781 records retrieved through the database searches and an additional 11 technical reports identified and included. After duplicates were removed, 18,295 records remained. 17,890 records were excluded after title and abstract screening, with 405 records remaining for full text review. Of these, 113 studies were excluded due to not meeting pre-specified inclusion criteria. There were 292 studies that reported some form of HIV incidence data (e.g., cumulative incidence, number of seroconversions, incidence rate), and 237 for which at least one incidence rate estimate was reported or could be derived (n = 229 reported and n = 8 derived, Figure 1; Supplemental Table 1).

**Figure 1:**
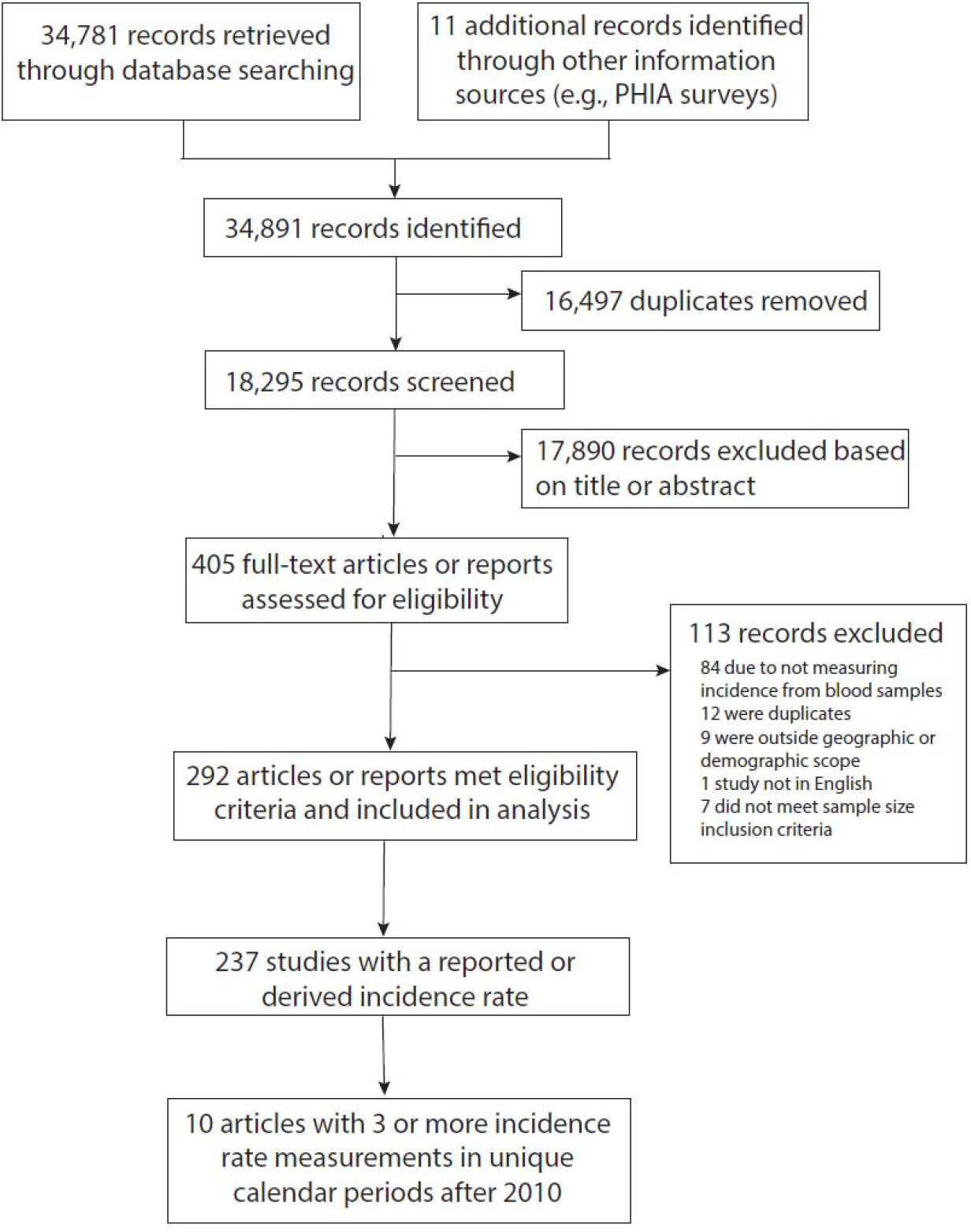
Flow of papers from literature search to full-text review. 34,781 papers published between 2010-2019 were identified through PubMed, Embase, Scopus, and OVID global health and 11 additional papers were identified through alternative sources including PHIA surveys. 16,497 duplicates were removed using Covidence. Of the 18,295 studies screened, 292 papers met eligibility criteria and were included in the analysis.

The 292 studies reported incidence data from 23 sub-Saharan countries (Figure 2A). Most studies were conducted in South Africa (n = 102), Uganda (n = 46), Kenya (n = 41), Zimbabwe (n = 21) and Tanzania (n = 14). Of note, there were 43 incidence estimates reported from multi-country studies (e.g. studies reporting combined estimates from two or more countries). While the number of studies published per country correlated with total case burden (Figure 2B), there were 26 countries in sub-Saharan Africa with no published incidence data. The majority of these countries were in western and central Africa, including Angola, Cape Verde, Central African Republic, Chad, Congo, Democratic Republic of Congo, Djibouti, Equatorial Guinea, Eritrea, Gabon, Gambia, Guinea, Liberia, Madagascar, Mauritius, Sierra Leone, Senegal, South Sudan, and Togo.

**Figure 2:**
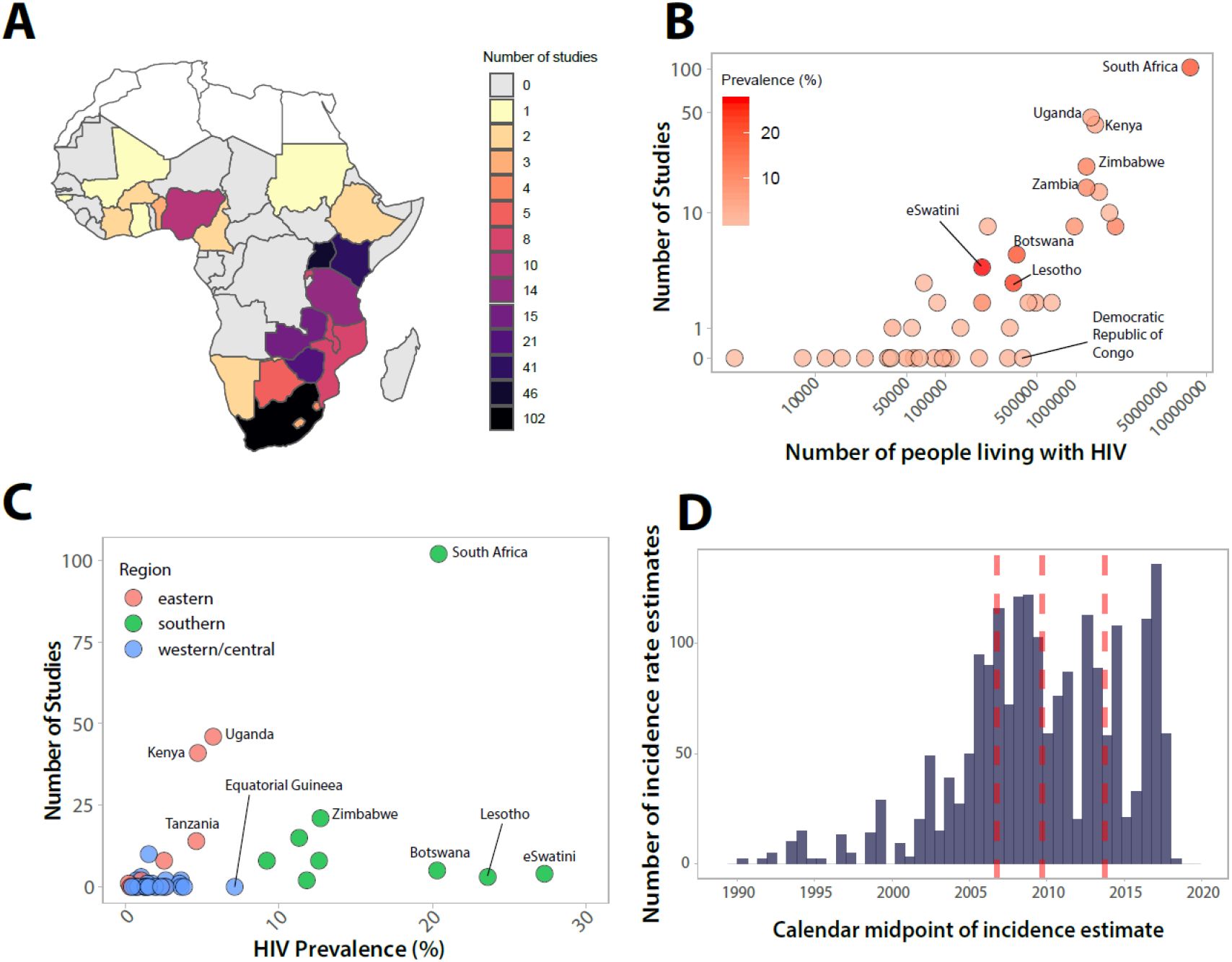
Number of studies reporting empirical HIV incidence data published between 2010-2019 by location, HIV burden, and study period. (A). Number of studies by country in sub-Saharan Africa. Studies only reporting incidence data across countries (e.g. South Africa and Zambia combined) were not included. (B) Number of studies by number of people living with HIV. Number of people living with HIV taken as the mean estimated number of adults (15+) living with HIV in 2018 from UNAIDS. (C) Number of studies by HIV prevalence and region. Prevalence taken as the 2018 adult (15-49) prevalence estimate from UNAIDS. (D) Distribution of calendar midpoints for HIV incidence rate estimates. The median and interquartile range of calendar midpoints are shown in red.

Figure 2C shows the number of studies reporting incidence data versus the prevalence of infection at the country level using 2018 national prevalence estimates from USAID^12^. Of the five countries with the highest HIV prevalence, there were only two with more than 10 studies published since 2010 (South Africa with 102 estimates and an HIV prevalence of 20.4%, Zimbabwe with 21 estimates and a prevalence of 12.7%). The remaining three countries, eSwatini, Lesotho, and Botswana had prevalences of 27.3%, 23.6% and 20.3%, but only had 4, 3, and 5, incidence studies published, respectively (Figure 3A).

**Figure 3:**
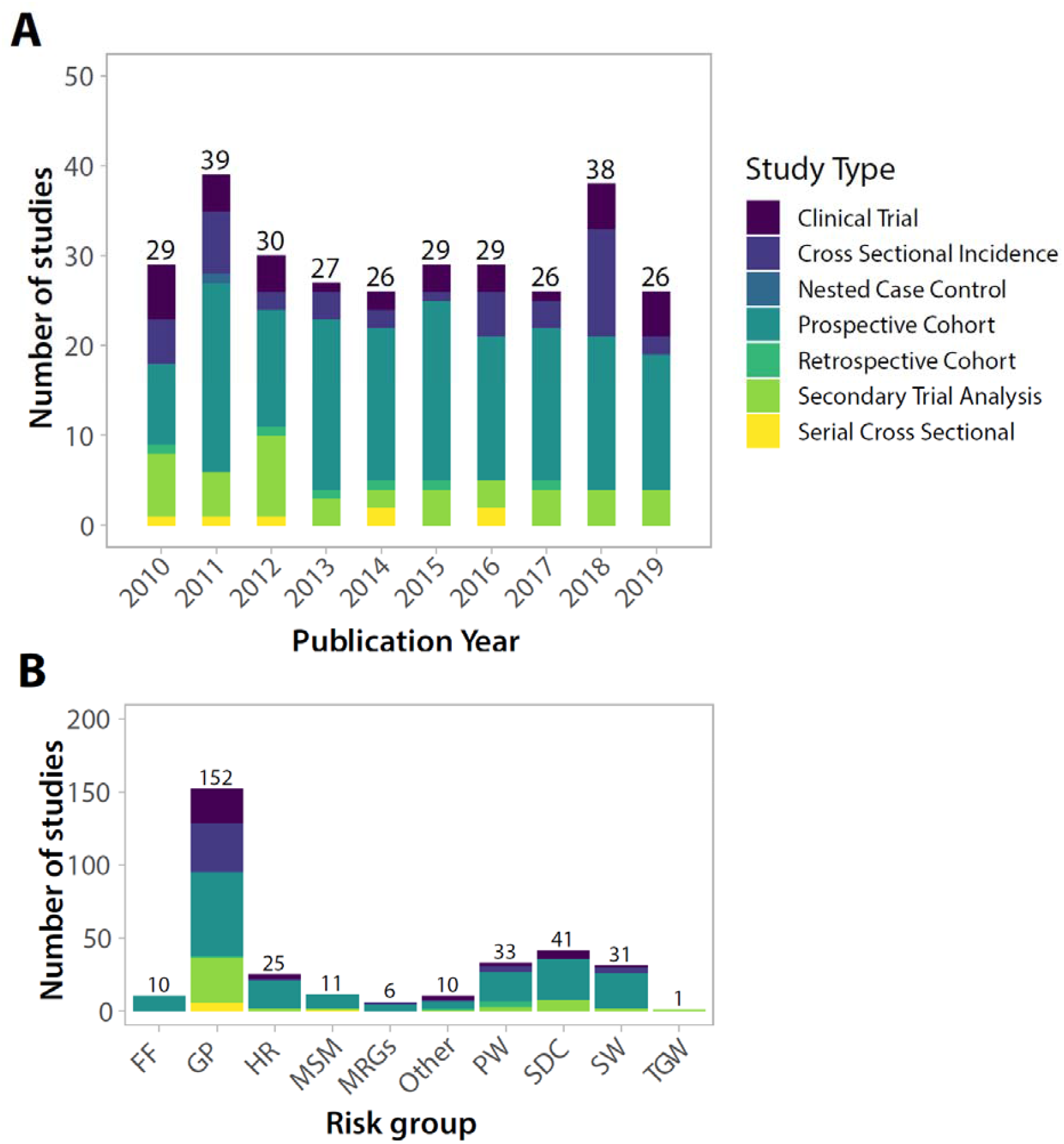
Number of studies reporting empirical HIV incidence data by year, and risk group. (A) Number of studies published by calendar year broken down by study type (B) Number of studies published by risk group broken down by study type. FF=fisherfolk, GP=general population; HR=high risk (e.g., women who report multiple partners); MSM=men who have sex with men; MSRGs=multiple risk groups (e.g., SW and MSM); PW=pregnant women; SDC=serodiscordant couples; SW=sex workers; TGW= transgender women

Publication of incidence data fluctuated little over calendar time (Figure 3A). Most incidence estimates were derived from prospective observational cohort studies (n=163), followed by secondary analyses of RCTs (n = 46), cross-sectional incidence studies (n = 42), primary analyses of RCTs (n=34), serial cross-sectional studies (n = 7), retrospective cohort studies (n = 6), and nested case control studies (n = 1). Of note, there were 7 studies reporting incidence data from two design classifications. Figure 3B shows studies disaggregated by population risk group. 152 estimates were drawn from GP studies, 41 from SDC, 33 from PW, 31 from sex workers SW, 11 from MSM, and 10 from FF communities in eastern Africa.

Next we summarized HIV incidence trends over calendar time for GP studies by sex and geographic region (Figure 4). A total of 50 studies contributed 146 estimates (n=83 for women; n=63 for men) to the analysis. Across both southern and eastern Africa, we observed declines in HIV incidence since 2010 in both sexes. In eastern Africa, annual incidence declined by 0.16/1000 pys (95%CI: 0.06-0.26; p=0.004) among men and by 0.16/1000 pys (95%CI: −0.01-0.33; p=0.060) among women. HIV incidence fell more rapidly in southern Africa, declining by 0.25/1000 pys (95%CI: 0.17-0.34; p<0.0001) annually among men and by 0.42/1000 pys (95%CI: 0.23-0.62; p=0.0002) annually among women. In sensitivity analyses, we excluded studies that did not report either an upper or lower age range (n=15) and observed similar trends (Supplemental Figure 1).

**Figure 4:**
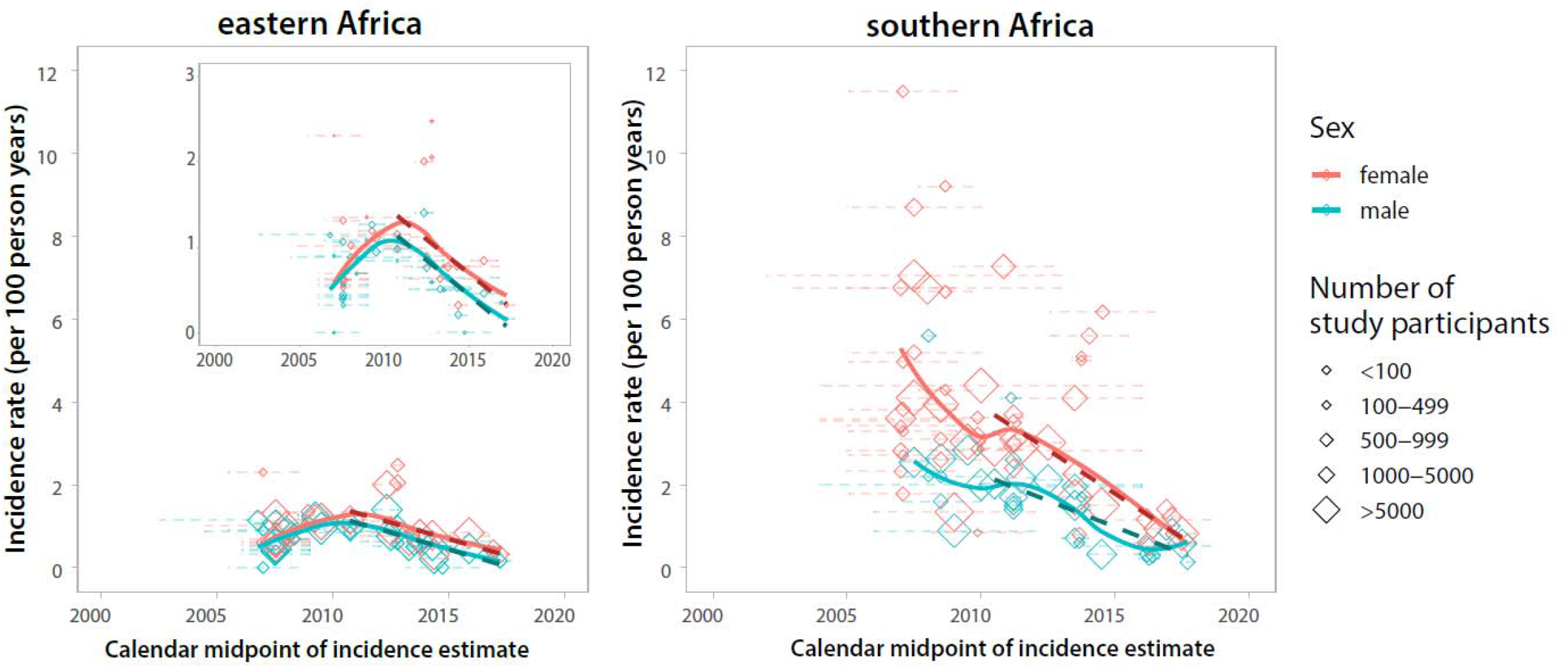
HIV Incidence rates in general population studies over time by sex for eastern and southern Africa. We included incidence rate estimates from general population studies in southern and eastern Africa that had a study mid-point from 2007 onward (this corresponds to the 25^th^ percentile of all calendar mid-points reported, Figure 2D). Diamonds represent the calendar midpoint of the incidence rate estimate while error bars represent the start and end date of the time interval over which the incidence rate was measured. Estimates are only shown for studies with an age range spanning 26 years or greater (e.g. an HIV estimate for individuals 18 to 44 years). Dashed lines show incidence trends fit using linear regression. Solid lines represented smoothed curves fit using loess regression. An inset is included for eastern Africa to highly trends with a y-axis restricted to 3 per 100 person-years.

We also examined HIV incidence trends within studies that had more than three incidence rate measurements in unique calendar periods since January 1, 2010 (Figure 5). Of the 292 studies, only 38 (13%) studies reported incidence rate estimates for more than one unique calendar period (Supplemental Figure 2), and of these studies only 10 had 3 or more measurements since 2010. Within nine of these studies, a decline in HIV incidence was observed over calendar time. The single outlier study showed rising incidence among pregnant women in South Africa^13^.

**Figure 5:**
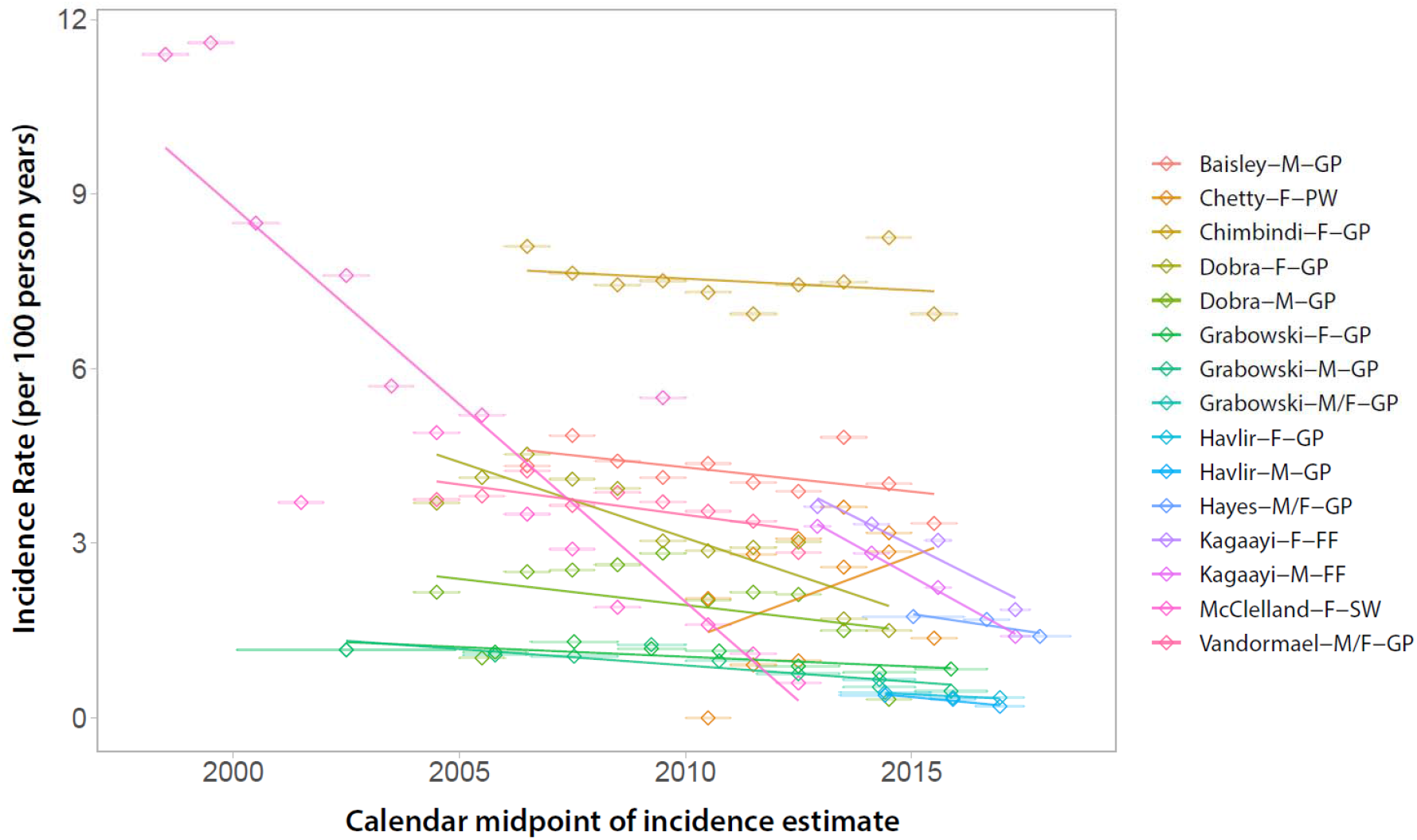
HIV incidence trends from 10 studies with three or more incidence rate measurements after 2010. Data is shown overall and disaggregated by sex where possible. The legend shows the first author, sex (M=male; F=female), and risk group (GP=General population; PW=pregnant women; FF=fisherfolk; SW=sex worker).

Figures 6 summarizes the most recent HIV incidence estimates since 2010 for GP studies in southern Africa. National estimates ranged from 0.37 per 100 pys in Malawi (2015-2016) to 2.4 per 100 pys in eSwatini (2010-2011). However, by 2016-2017, HIV incidence declined to 1.13 per 100 pys in eSwatini. There was substantial variation across national and sub-national incidence estimates in South Africa with rates ranging 0.69 at the national level to 3.28 pys sub-nationally. Of the 4 measurements exceeding 2 per 100 pys, 3 were in South Africa and 1 in eSwatini. Overall, South Africa reported the highest rates of HIV incidence in the general population, and these estimates were predominately concentrated in KwaZulu-Natal, a coastal province in the southeast.

**Figure 6.**
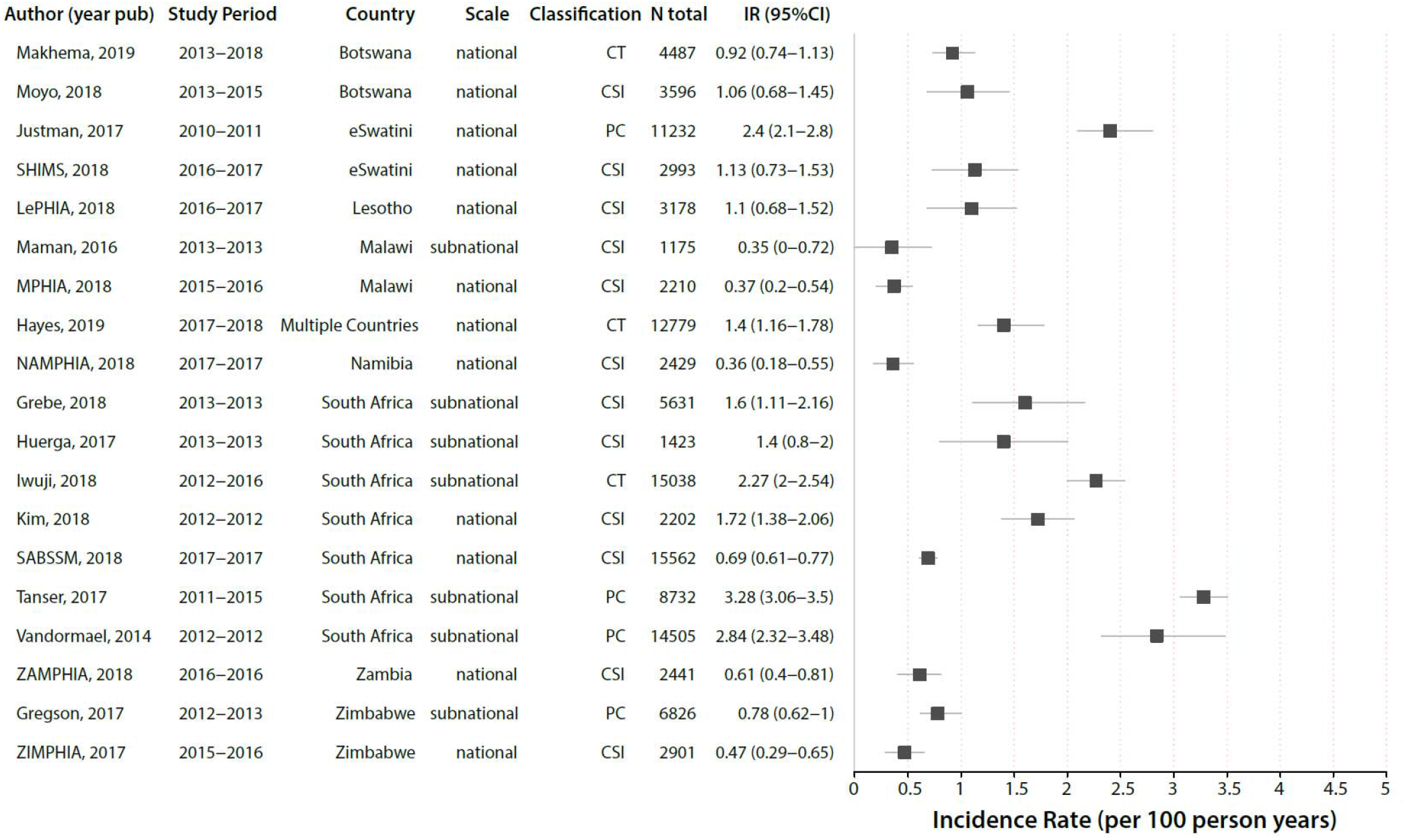
Forest plot of HIV incidence estimates after 2010 for general population studies in southern Africa. Only the most recent HIV incidence estimate for a cohort/study population are shown. Incidence rates are reported as the number of new cases per 100 person-years and the error bars represent 95% CI. Study references are reported in supplemental Table 1. CT=clinical trial; CSI=cross-sectional incidence study; PC=prospective cohort

Supplemental Figure 3 shows the most recent HIV incidence estimates since 2010 for general population studies in eastern Africa. Estimates ranged from 0.06 per 100 pys (95%CI: 0-0.12) in Ethiopia to 3.4 per 100 pys (no confidence interval reported) in Sudan. National studies were conducted in Ethiopia, Kenya, Rwanda, Uganda, and Tanzania and all HIV incidence estimates were below 1 per 100 pys. There were only two general population studies in west Africa since 2010, including the Cameroon PHIA in 2017-18 which reported an HIV incidence rate of 0.27 (95%CI: 0.14-0.41) per 100 pys^14^, and another study in blood donors in Cote d’Ivoire (0.07 per 100 pys; 95%CI: 0.06-0.08)^15^.

We also collated recent HIV incidence estimates for key populations, including SW, MSM, and FF (Supplementary Figures 4-6). Overall, there were few incidence estimates available for these groups since 2010; however, incidence rates in these populations were typically much higher than that observed in GP studies. All estimates for SWs (n=7) were derived from cohort studies and ranged from 0.6 to 3.4 per 100 pys (Supplemental Figure 4). Incidence estimates among MSM cohorts (n=5) were highly variable ranging from 1.03 per 100 pys (95% CI: 0.03-5.79) in Kenya to 15.4 per 100 pys (95%CI: 12.3-19) in Nigeria (Supplemental Figure 5). Among FF populations, HIV incidence ranged from 1.59 to 6.04 per 100 pys (Supplemental Figure 6).

Lastly, we directly compared HIV incidence estimates between men and women within GP studies (Figure 7). Of the 237 studies with incidence rates reported, 108 (46%) reported incidence estimates for women only and 18 (8%) for men only. There were an additional 23 (10%) studies that reported combined estimates for men and women only, and 88 (37%) studies reported sex stratified results including both sexes. Among 44 GP studies reporting 96 sex stratified incidence estimates for age bins ≥26 years, female HIV incidence was higher in 78 (81%) instances and the median female to male HIV incidence ratio was 1.45 (IQR: 1.12-1.83) overall (Figure 7A). Temporal assessment of the female to male incidence rate ratio showed evidence for growing disparity by sex over calendar time, increasing by 2.4% per year on average (95CI:1.14-3.74%, p=0.0003; Figure 7B).

**Figure 7.**
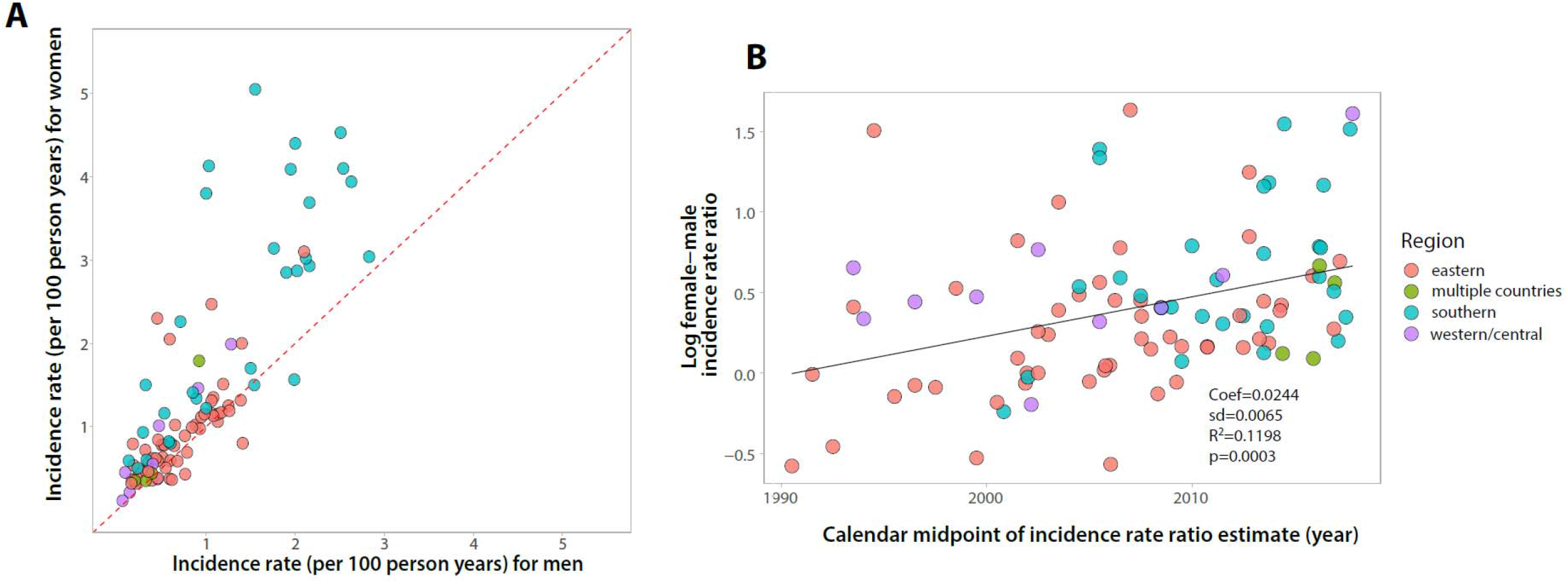
HIV incidence rates among men and women in general population studies. (A) Plot of male vs. female HIV incidence per 100 person-years. Dashed red line is the identify line; points above the line represent higher female HIV incidence while points below the line represent higher male HIV incidence. (B) Scatterplot of the log female-male HIV incidence ratio over calendar time with fitted line (solid black line) estimated using linear regression.

## DISCUSSION

Monitoring HIV incidence trends is critical for assessing progress and targeting resources. Our analysis of directly observed HIV incidence data demonstrates that the rate of new infections is rapidly declining in eastern and southern Africa among both men and women. These findings are consistent with UNAIDS incidence models which have also shown reductions in new cases continent-wide over the last decade^1,6^. However, there are notable gaps in the availability of empiric HIV incidence data, particularly for western and central Africa and among key populations, including MSM among whom HIV incidence was highest. HIV incidence remains higher among women, despite overall reductions in acquisition risk in both sexes, and the incidence gap between sexes appears to be growing with time.

The last comprehensive systematic review of the empiric HIV incidence literature in sub-Saharan Africa was published in 2009 and included 57 studies conducted in 14 sub-Saharan African countries^9^. In this this systematic review of incidence data published between 2010 and 2019, incidence estimates were available for 23 sub-countries; however, data were non-existent or sparse in many countries. Of the 292 studies included in this study, only 20 (7%) were conducted in western and central Africa, despite this region accounting for an estimated 25% of all incidence cases in sub-Saharan Africa in 2019 ^1^. Critically, 16 countries in western and central Africa reported no incidence data at all, illuminating critical geographic gaps in surveillance and interventional research efforts. While the number of studies conducted per country tended to trend with higher HIV case burden and prevalence, there were several countries with either moderately high case burden or prevalence reporting no or very limited data, including the Democratic Republic of Congo and Equatorial Guinea. Similar geographic disparities in incidence data have been reported earlier^9^.

We observed steadily declining HIV incidence rates across general population studies in eastern and southern Africa since 2010. While HIV incidence was consistently higher among women than men, there were significant reductions in incidence rates in both sexes. Notably, there was a 50% reduction in HIV incidence in eSwatini between 2010 and 2017, the largest national decline reported. These broad reductions in HIV incidence are likely the partial result of massive global efforts to rollout and scale-up HIV prevention and treatment interventions, including antiretroviral therapy (ART) and voluntary medical male circumcision (VMMC). Indeed, several population-based cohorts and RCTs have reported on the link between ART and VMMC and declines in population-level incidence^16–22^.

Overall, only 16% of studies reported HIV incidence measurements at multiple time points within the same study population. However, nearly half of these studies ended prior to 2010 before the scale-up of major prevention and treatment interventions, resulting in unfortunate lost opportunity to evaluate their impact. Of the ten studies that obtained more than two incidence measurements after 2010, all but one showed evidence of declining HIV incidence, consistent with our analyses of incidence across general population studies. Of note, two of these nine studies were conducted in high risk key populations, one among Lake Victoria fisherfolk and the other among sex workers^16,23^, suggesting incidence reductions may not be exclusive to general population cohorts.

At the sub-national level we found substantial variation in HIV incidences rates. These findings are consistent with recent a study that reported marked small-scale spatial heterogeneities in disease burden across the continent^24^. While some variation in rates is likely due to temporal heterogeneities, there were countries that reported estimates over similar calendar periods (e.g., South Africa) with substantial variation in incidence. Prior studies in South Africa have identified high risk corridors of transmission with elevated HIV incidence rates compared to the surrounding population^25^. Furthermore, these areas tended to correlate with higher prevalence, suggesting that current maps of HIV prevalence, potentially in combination with uptake of HIV prevention and treatment data, could be used to strategically implement HIV programs and target interventions, such as pre-exposure prophylaxis (i.e., PrEP), to high risk populations.

In general population studies, HIV incidence was nearly always higher among women, a finding consistent with a recent systematic review of incidence among African adolescents and young adults 15-24 years old^26^. Notably, nearly half of all incidence studies were conducted among women, while just under 10% were conducted among men only. A better understanding of incidence and associated risk factors for transmission among men might potentially result in lower female transmission given the large preponderance of heterosexual HIV transmission in Africa^1^. Studies throughout Africa show lower uptake of HIV prevention and treatment interventions among men, but also lower participation rates in many HIV incidence studies^27^. We also found the ratio of female to male HIV incidence increased over calendar time. It is unclear what is driving this growing HIV disparity between men and women; however, lower male susceptibility due to increasing VMMC coverage and higher female ART uptake may partially explain this observation.

In line with prior systematic reviews^28,29^, we found substantially higher estimates of HIV incidence among key populations, including Lake Victoria fisherfolk, MSM, and sex workers, as compared to general population cohorts. However, there were relatively few studies in these populations overall, and among them there was substantial variation in HIV incidence. MSM had the highest HIV incidence rates of any sub-group, consistent with prior studies showing extremely high HIV prevalence among African MSM^28^.

There are limitations to this study. First, the methods used to obtain HIV incidence estimates across studies, including study design, testing algorithms, and definitions of incidence were highly variable. Second, upper and lower age eligibility criteria were often unreported. In the case of general population studies that did not report an upper or lower age bound, we imputed the range to be 15-65 years. In a sensitivity analysis assessing incidence trends over calendar time, we excluded these studies and found no qualitative differences in our inferences. Third, there were many countries and geographic locations for which there were no incidence data available. It is unclear whether the data presented here are representative of empirical trends in unobserved locations, most notably western and central Africa. Reassuringly, our results are consistent with UNAIDS regional models of incidence trends also showing declines in new cases over the same time frame^1^. Additionally, our analysis of incidence trends were only done among general population cohorts and thus may not reflect changes among key populations. Fourth, published data may not be fully representative of all incidence data, particularly studies with higher incidence rates may be more likely to be published than those with low or zero incidence. Lastly, the results of this study may have been impacted by our own review methodologies, including the search strategy and our inclusion and exclusion criteria. Critically, we excluded studies that were not in English, which may explain some of the data gaps observed in western and central Africa.

In conclusion, our findings provide empirical evidence that HIV incidence in eastern and southern Africa has declined over the last decade. The widespread scale-up and uptake of HIV treatment and prevention interventions has been linked to incidence declines within individual studies previously^16–22^, and are likely contributing to the broad scale incidence reductions observed here. Sustained commitment to these programs and new investment in innovative technologies and approaches may ultimately help achieve HIV elimination goals in sub-Saharan Africa. However, future successes remain in the balance as global funding for HIV programs in low and middle-income countries dwindles^2^. The COVID-19 pandemic presents further challenges as critical resources are diverted and mobility restrictions impact uptake of interventions, in particular, HIV testing and timely initiation of ART^30,31^. Ongoing surveillance, including addressing geographic and population disparities in HIV incidence data, will be critical to maintaining and building upon past success in the coming decade.

## Data Availability

All underlying data and code is available upon request. Data will be made publicly available following publication.

## Acknowledgements

This work was supported by the National Institute of Allergy and Infectious Diseases (K01AI125086 to M.K.G. and R01AI120938 and R01AI128779 to A.A.R.T).

We thank Dr. Jeff Eaton if his thoughtful comments and advice on this manuscript.

