## Supplemental Figures for "Declining HIV incidence in sub-Saharan Africa: a systematic review and meta-analysis of empiric data"

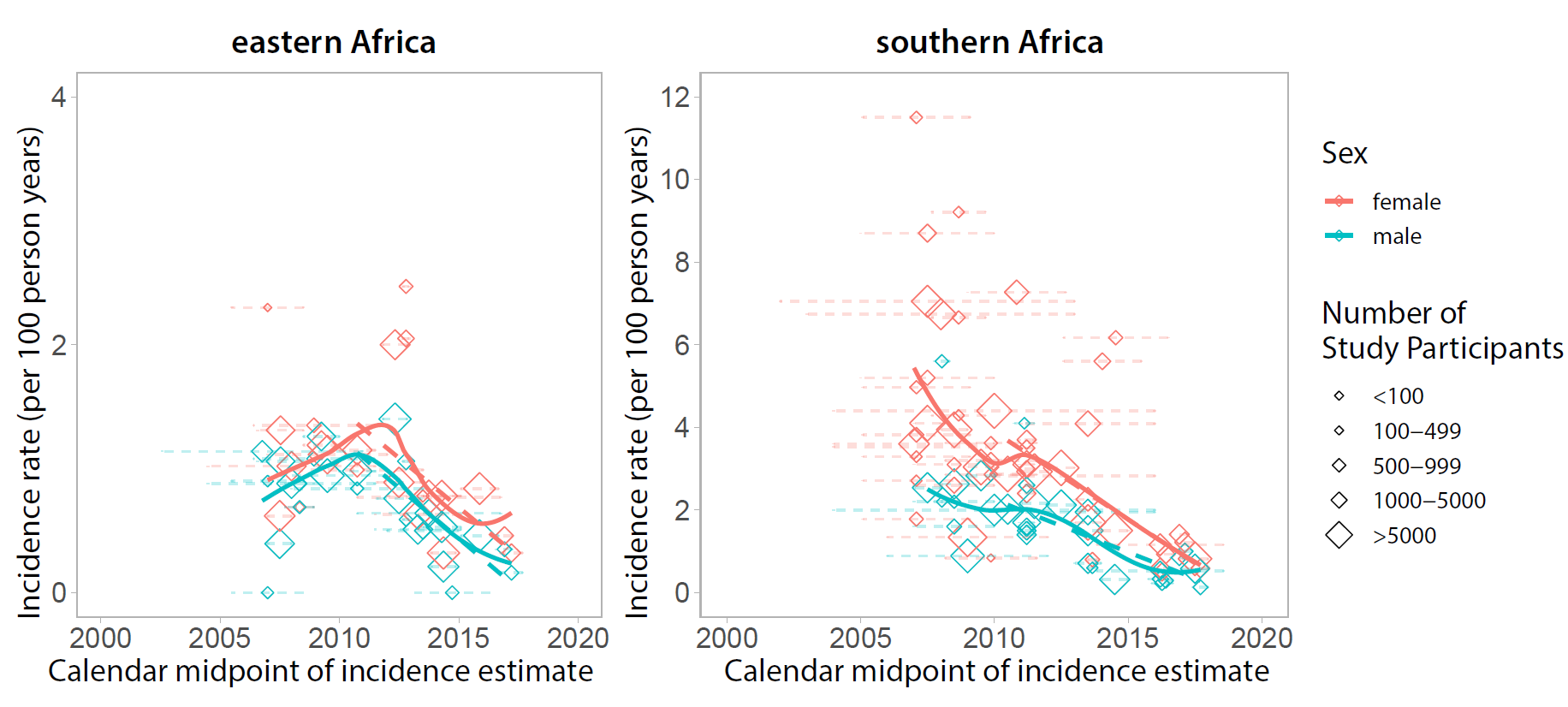

**Supplemental Figure 1**. Sensivity analysis of time trends excluding studies that did not report a minimum or maximum age range for the study.

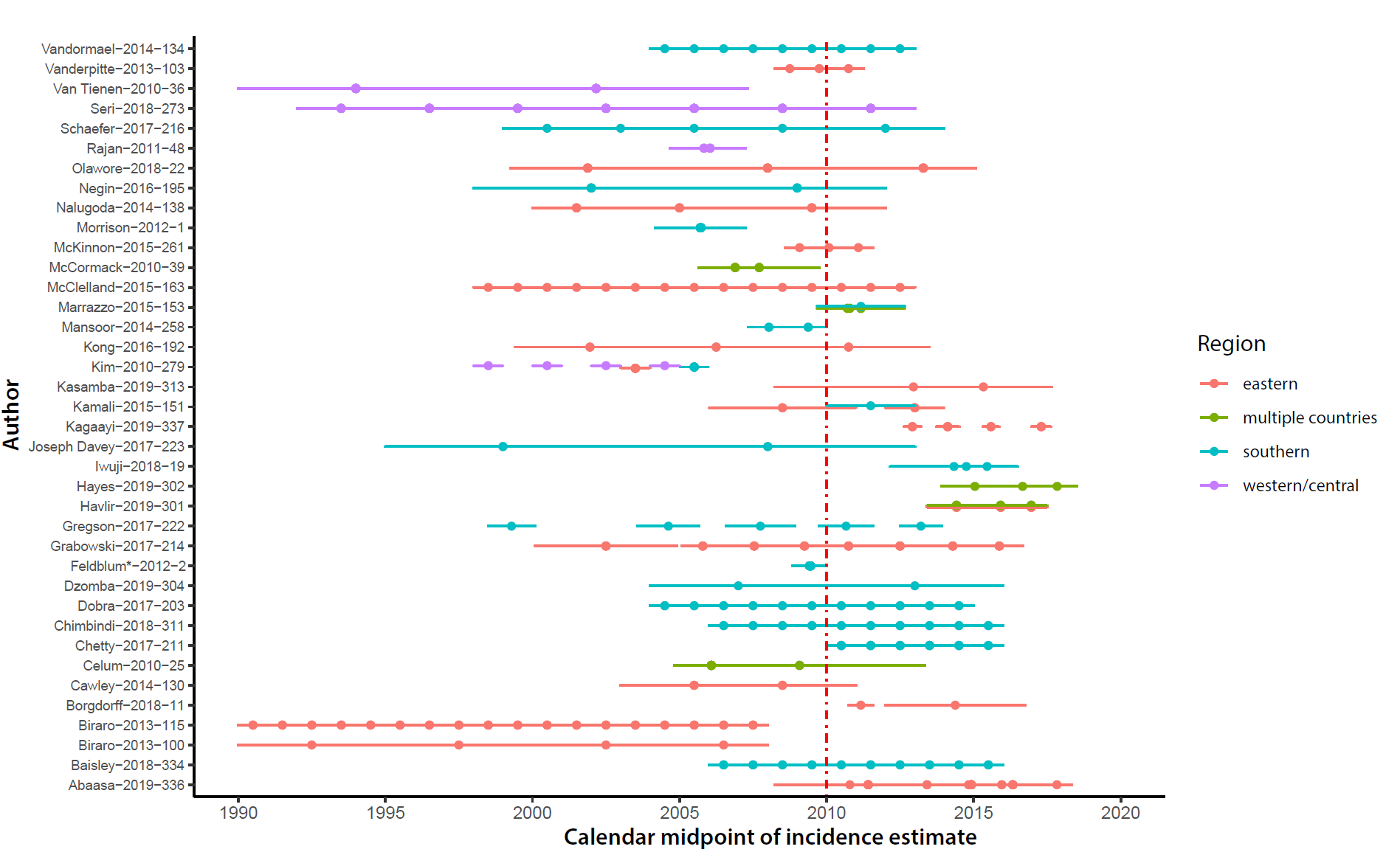

**Supplemental Figure 2. 38 studies reporting two or more incidence rate estimates in unique calendar periods**. The dots represent the calendar midpoint of the incidence estimate and the lines the time period over which incidence was measured. *This study’s prior incidence estimate was in 1993 and is not shown.

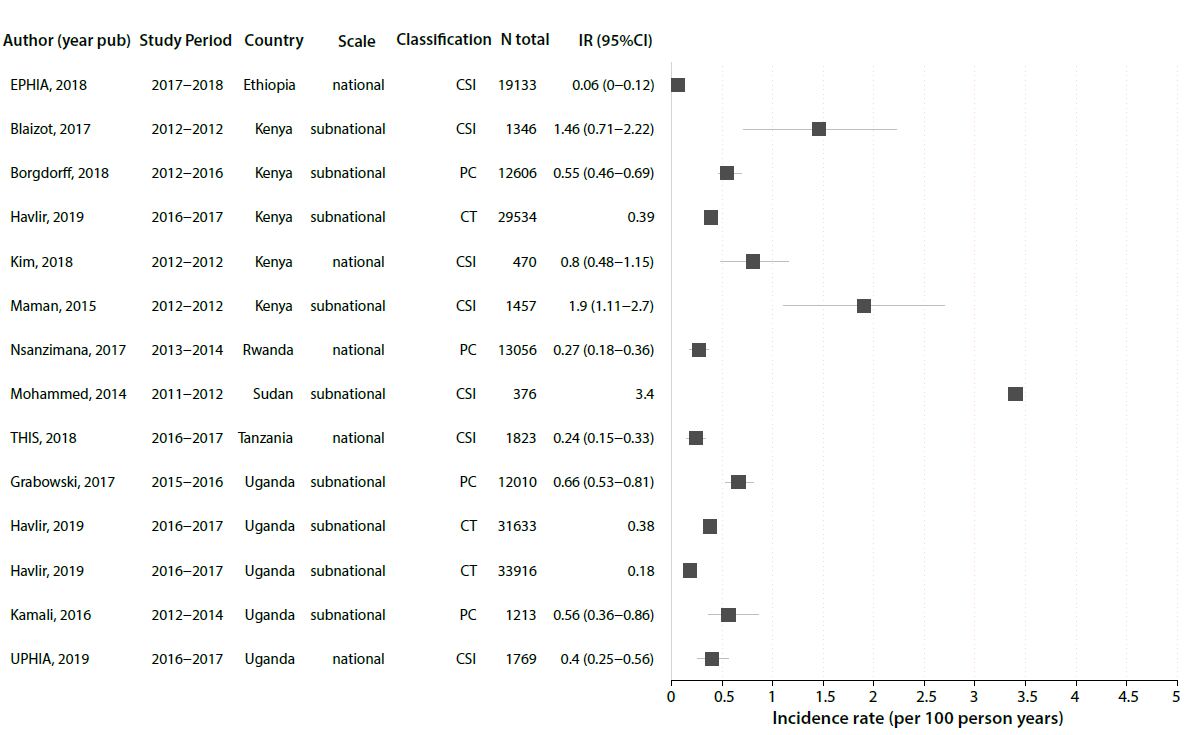

**Supplemental Figure 3. Forest plot of HIV incidence estimates after 2010 for general population studies in eastern Africa**. Only the most recent HIV incidence estimate for a cohort/study population are shown. Incidence rates are reported as the number of new cases per 100 person-years and the error bars represent 95% CI. Estimates without error bars did not report a confidence interval/standard error for the estimate. Study references are reported in supplemental Table 1. PC=prospective cohort; CSI=cross-sectional incidence study; CT=Clinical trial

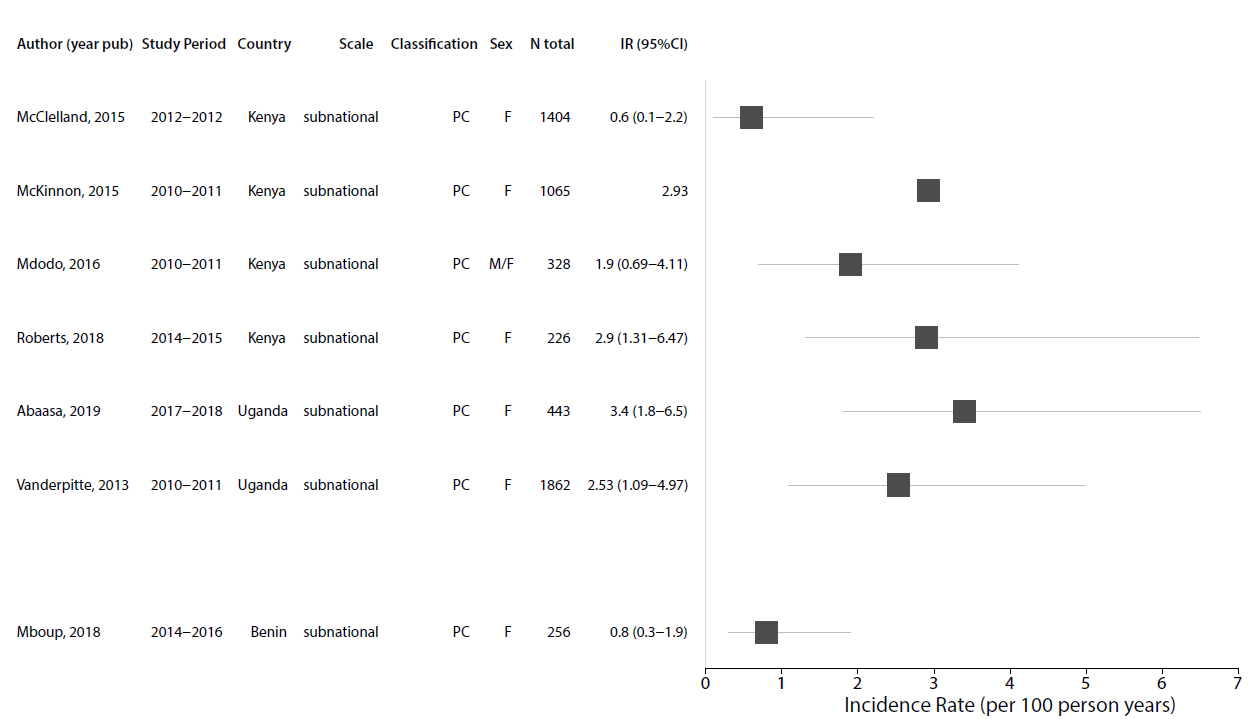

**Supplemental Figure 4. Forest plot of HIV incidence estimates after 2010 for studies among female sex workers (SW)**. Only the most recent HIV incidence estimate for a cohort/study population are shown. Incidence rates are reported as the number of new cases per 100 person-years and the error bars represent 95% CI. Estimates without error bars did not report a confidence interval/standard error for the estimate. Study references are reported in supplemental Table 1. PC=prospective cohort; SCS=serial cross-sectional study

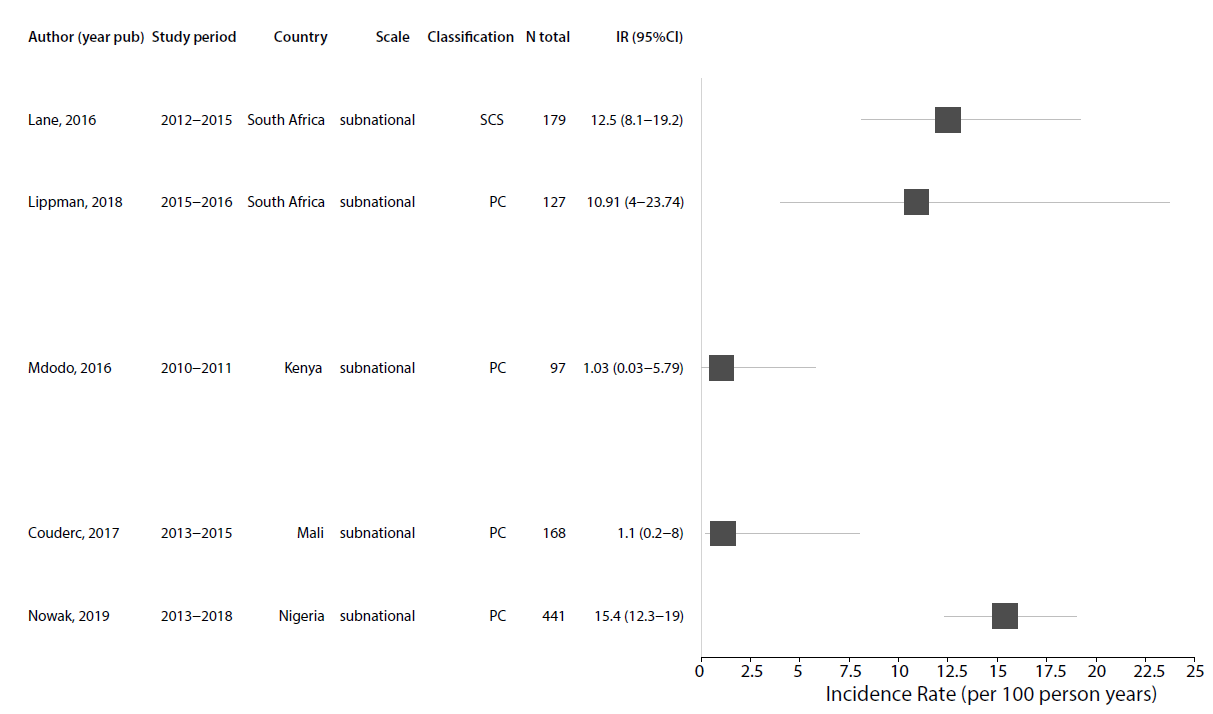

**Supplemental Figure 5. Forest plot of HIV incidence estimates after 2010 for studies among men who have sex with men (MSM)**. Only the most recent HIV incidence estimate for a cohort/study population are shown. Incidence rates are reported as the number of new cases per 100 person-years and the error bars represent 95% CI. Estimates without error bars did not report a confidence interval/standard error for the estimate. Study references are reported in supplemental Table 1. PC=prospective cohort; SCS=serial cross-sectional study

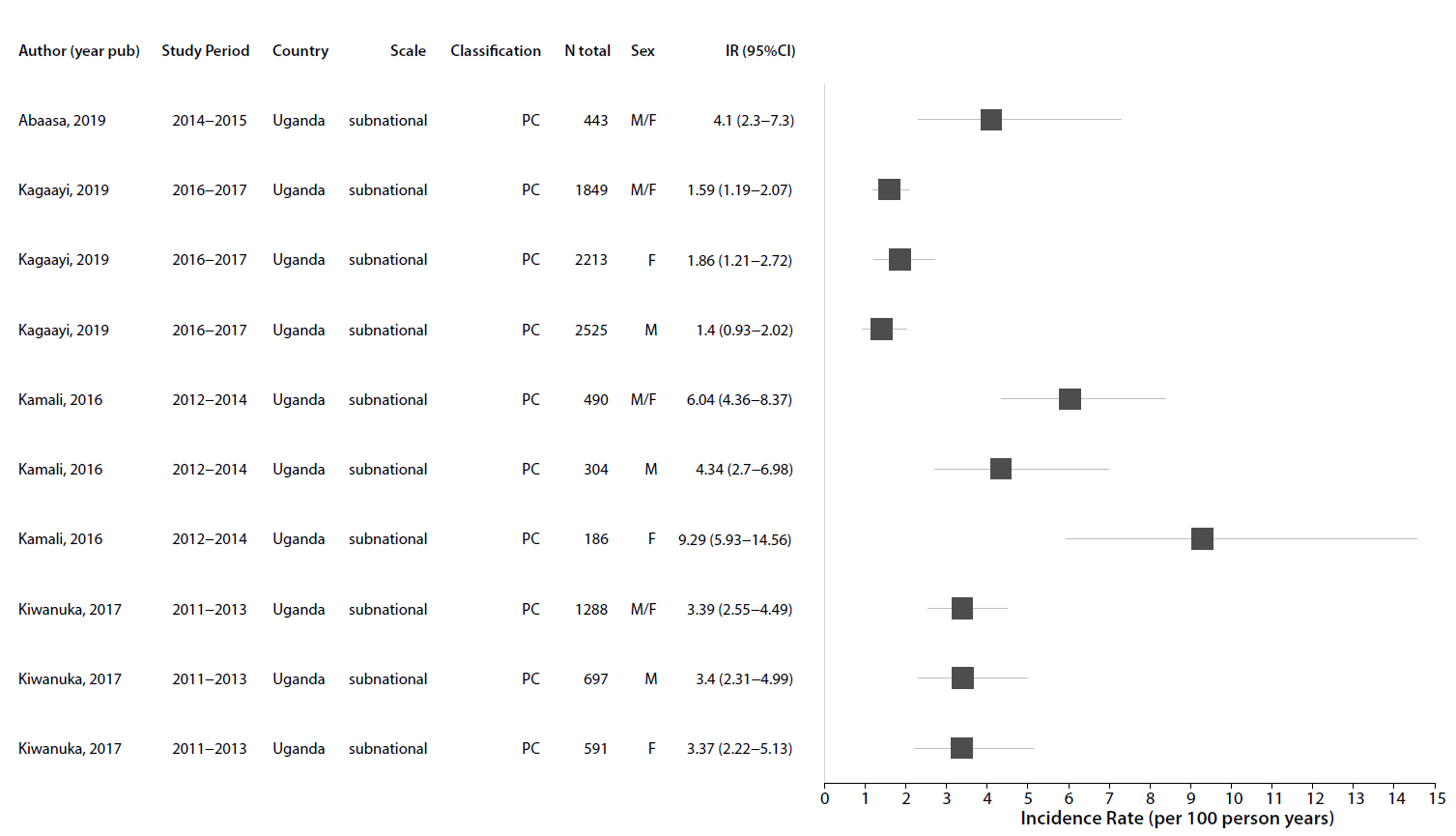

**Supplemental Figure 6. Forest plot of HIV incidence estimates after 2010 for studies of Lake Victoria fisherfolk in Eastern Africa**. Only the most recent HIV incidence estimate for a cohort/study population are shown. Incidence rates are reported as the number of new cases per 100 person-years and the error bars represent 95% CI. Estimates without error bars did not report a confidence interval/standard error for the estimate. Study references are reported in supplemental Table 1. PC=prospective cohort
