## Supplemental Tables for "Declining HIV incidence in sub-Saharan Africa: a systematic review and meta-analysis of empiric data"

**Supplemental Table 1:** Full list of the 292 studies reporting HIV incidence data

| **Paper ID** | **Title** | **Author** | **Year of Publication** | **DOI** | **Study Classification** | **High Risk Group** | **Location(s)** | **Region** |
| --- | --- | --- | --- | --- | --- | --- | --- | --- |
| 1 | Hormonal contraception and the risk of HIV acquisition among women in South Africa | Morrison | 2012 | 10.1097/QAD.0b013e32834fa13d | Secondary Trial Analysis | General Population | South Africa | Southern |
| 2 | HIV incidence and prevalence among cohorts of women with higher risk behaviour in Bloemfontein and Rustenburg, South Africa: a prospective study | Feldblum | 2012 | 10.1136/bmjopen-2011-000626 | Prospective Cohort | High Risk | South Africa | Southern |
| 3 | The relationship between age of coital debut and HIV seroprevalence among women in Durban, South Africa: a cohort study | Wand | 2012 | 10.1136/bmjopen-2011-000285 | Secondary Trial Analysis | General Population | South Africa | Southern |
| 4 | Bacterial Vaginosis Associated with Increased Risk of Female-to-Male HIV-1 Transmission: A Prospective Cohort Analysis among African Couples | Cohen | 2012 | 10.1371/journal.pmed.1001251 | Secondary Trial Analysis | Serodiscordant Couple | Multiple Countries | Multiple Countries |
| 5 | Identifying HIV infection in South African women: How does a fourth generation HIV rapid test perform? | Bhowan | 2012 | 10.4102/ajlm.v1i1.4 | Prospective Cohort | Pregnant Women | South Africa | Southern |
| 6 | Risk Factor Detection as a Metric of STARHS Performance for HIV Incidence Surveillance Among Female Sex Workers in Kigali, Rwanda | Braunstein | 2012 | 10.2174/1874613601206010112 | Prospective Cohort/Cross Sectional Incidence | Sex Worker | Rwanda | Eastern |
| 7 | Mother to child transmission of HIV among Zimbabwean women who seroconverted postnatally: prospective cohort study | Humphrey | 2010 | 10.1136/bmj.c6580 | Secondary Trial Analysis | Pregnant Women | Zimbabwe | Southern |
| 8 | Characteristics, medical management and outcomes of survivors of sexual gender-based violence, Nairobi, Kenya | Buard | 2013 | 10.5588/pha.13.0012 | Retrospective Cohort | Other | Kenya | Eastern |
| 9 | Efficacy of oral pre-exposure prophylaxis (PrEP) for HIV among women with abnormal vaginal microbiota: a post-hoc analysis of the randomised, placebo-controlled Partners PrEP Study | Heffron | 2017 | 10.1016/S2352-3018(17)30110-8 | Secondary Trial Analysis | Serodiscordant Couple | Multiple Countries | Multiple Countries |
| 10 | HIV incidence in western Kenya during scale-up of antiretroviral therapy and voluntary medical male circumcision: a population-based cohort analysis | Borgdorff | 2018 | 10.1016/S2352-3018(18)30025-0 | Prospective Cohort | General Population | Kenya | Eastern |
| 11 | Gonorrhea, Chlamydia and HIV incidence among female sex workers in Cotonou, Benin: A longitudinal study | Diabate | 2018 | 10.1371/journal.pone.0197251 | Prospective Cohort | Sex Worker | Benin | Western/Central |
| 12 | Incidence of sexually transmitted infections during pregnancy | Teasdale | 2018 | 10.1371/journal.pone.0197696 | Secondary Trial Analysis | Multiple Risk Groups | Multiple Countries | Multiple Countries |
| 13 | Effects of injectable progestogen contraception versus the copper intrauterine device on HIV acquisition: sub-study of a pragmatic randomised controlled trial | Hofmeyr | 2017 | 10.1136/jfprhc-2016-101607 | Clinical Trial | General Population | South Africa | Southern |
| 14 | Household survey of HIV incidence in Rwanda: a national observational cohort study | Nsanzimana | 2017 | 10.1016/S2352-3018(17)30124-8 | Prospective Cohort | General Population | Rwanda | Eastern |
| 15 | Effect of population viral load on prospective HIV incidence in a hyperendemic rural African community | Tanser | 2017 | 10.1126/scitranslmed.aam8012 | Prospective Cohort | General Population | South Africa | Southern |
| 16 | School Support as Structural HIV Prevention for Adolescent Orphans in Western Kenya | Cho | 2018 | 10.1016/j.jadohealth.2017.07.015 | Clinical Trial | Other | Kenya | Eastern |
| 17 | Detection and treatment of Fiebig stage I HIV-1 infection in young at-risk women in South Africa: a prospective cohort study | Dong | 2018 | 10.1016/S2352-3018(17)30146-7 | Prospective Cohort | High Risk | South Africa | Southern |
| 18 | Universal test and treat and the HIV epidemic in rural South Africa: a phase 4, open-label, community cluster randomised trial | Iwuji | 2018 | 10.1016/S2352-3018(17)30205-9 | Clinical Trial | General Population | South Africa | Southern |
| 19 | Impact of early antiretroviral therapy eligibility on HIV acquisition: household-level evidence from rural South Africa | Oldenburg | 2018 | 10.1097/QAD.0000000000001737 | Prospective Cohort | Other | South Africa | Southern |
| 20 | Bacterial vaginosis modifies the association between hormonal contraception and HIV acquisition | Haddad | 2018 | 10.1097/QAD.0000000000001741 | Prospective Cohort | Serodiscordant Couple | Zambia | Southern |
| 21 | Migration and risk of HIV acquisition in Rakai, Uganda: a population-based cohort study | Olawore | 2018 | 10.1016/S2352-3018(18)30009-2 | Prospective Cohort | General Population | Uganda | Eastern |
| 22 | Mother to child transmission of HIV among Zimbabwean women who seroconverted postnatally: prospective cohort study | Humphrey | 2010 | 10.1136/bmj.c6580 | Prospective Cohort | Pregnant Women | Zimbabwe | Southern |
| 23 | Intimate partner violence, relationship power inequity, and incidence of HIV infection in young women in South Africa: a cohort study | Jewkes | 2010 | 10.1016/S0140-6736(10)60548-X | Secondary Trial Analysis | General Population | South Africa | Southern |
| 24 | Acyclovir and Transmission of HIV-1 from Persons Infected with HIV-1 and HSV-2 | Celum | 2010 | 10.1056/NEJMoa0904849 | Clinical Trial | Serodiscordant Couple | Multiple Countries | Multiple Countries |
| 25 | Behavioral Changes Associated With Testing HIV-Positive Among Sexually Transmitted Infection Clinic Patients In Cape Town, South Africa | Kalichman | 2010 | 10.2105/AJPH.2009.162602 | Prospective Cohort | High Risk | South Africa | Southern |
| 26 | Uptake of provider-initiated HIV testing and counseling among women attending an urban sexually transmitted disease clinic in South Africa – missed opportunities for early diagnosis of HIV infection | Kharsany | 2010 | 10.1080/09540120903254005 | Cross Sectional Incidence | High Risk | South Africa | Southern |
| 27 | Can HIV incidence testing be used for evaluating HIV intervention programs? A reanalysis of the Orange Farm male circumcision trial (ANRS-1265) | Fiamma | 2010 | 10.1186/1471-2334-10-137 | Secondary Trial Analysis/Cross Sectional Incidence | General Population | South Africa | Southern |
| 28 | Acute HIV Infections among Men with Genital Ulcer Disease in South Africa | Paz Bailey | 2010 | 10.1086/652785 | Secondary Trial Analysis | Other | South Africa | Southern |
| 29 | Identification of recent HIV infections and of factors associated with virus acquisition among pregnant women in 2004 and 2006 in Swaziland | Bernasconi | 2010 | 10.1016/j.jcv.2010.04.010 | Cross Sectional Incidence | Pregnant Women | eSwatini | Southern |
| 30 | Effectiveness and safety of tenofovir gel, an antiretroviral microbicide, for the prevention of HIV infection in women | Karim | 2010 | 10.1126/science.1193748 | Clinical Trial | General Population | South Africa | Southern |
| 31 | HIV acquisition is associated with prior high-risk human papillomavirus infection among high-risk women in Rwanda | Veldhuijzen | 2010 | 10.1097/QAD.0b013e32833cbb71 | Prospective Cohort | Sex Worker* | Rwanda | Eastern |
| 32 | Are Women Who Work in Bars, Guesthouses and Similar Facilities a Suitable Study Population for Vaginal Microbicide Trials in Africa? | Vallely | 2010 | 10.1371/journal.pone.0010661 | Prospective Cohort | High Risk | Tanzania | Eastern |
| 33 | Acute HIV infection among pregnant women in Malawi | Gay | 2010 | 10.1016/j.diagmicrobio.2009.12.001 | Prospective Cohort | Pregnant Women | Malawi | Southern |
| 34 | Pregnancy, Contraceptive Use, and HIV Acquisition in HPTN 039: Relevance for HIV Prevention Trials Among African Women | Reid | 2010 | 10.1097/QAI.0b013e3181bc4869 | Secondary Trial Analysis | High Risk | South Africa, Zambia, Zimbabwe | Multiple Countries |
| 35 | Two distinct epidemics: the rise of HIV-1 and decline of HIV-2 infection between 1990 and 2007 in rural Guinea-Bissau | Van Tienen | 2010 | 10.1097/QAI.0b013e3181bf1a25 | Serial Crosssectional Survey | General Population | Guinea-Bissau | Western/Central |
| 36 | HIV-1 Incidence Rates and Risk Factors in Agricultural Workers and Dependents in Rural Kenya: 36-Month Follow-Up of the Kericho HIV Cohort Study | Shaffer | 2010 | 10.1097/QAI.0b013e3181bcdae0 | Prospective Cohort | Other | Kenya | Eastern |
| 37 | Cofactors for HIV-1 Incidence during Pregnancy and Postpartum Period | Kinuthia | 2010 | NA | Retrospective Cohort | Pregnant Women | Kenya | Eastern |
| 38 | PRO2000 vaginal gel for prevention of HIV-1 infection (Microbicides Development Programme 301): a phase 3, randomised, double-blind, parallel-group trial | McCormack | 2010 | 10.1016/S0140-6736(10)61086-0 | Clinical Trial | General Population | Multiple Countries | Multiple Countries |
| 39 | Safety and Immunogenicity Study of Multiclade HIV-1 Adenoviral Vector Vaccine Alone or as Boost following a Multiclade HIV-1 DNA Vaccine in Africa | Jaoko | 2010 | 10.1371/journal.pone.0012873 | Clinical Trial | General Population | Multiple Countries | Multiple Countries |
| 40 | Associations between childhood adversity and depression, substance abuse and HIV and HSV2 incident infections in rural South African youth | Jewkes | 2010 | 10.1016/j.chiabu.2010.05.002 | Secondary Trial Analysis | General Population | South Africa | Southern |
| 41 | Screening for ‘window‐period’ acute HIV infection among pregnant women in rural South Africa | Kharsany | 2010 | 10.1111/j.1468-1293.2010.00838.x | Cross Sectional Incidence | Pregnant Women | South Africa | Southern |
| 42 | The incidence of HIV among women recruited during late pregnancy and followed up for six years after childbirth in Zimbabwe | Munjoma | 2010 | 10.1186/1471-2458-10-668 | Prospective Cohort | Pregnant Women | Zimbabwe | Southern |
| 43 | Expanded safety and acceptability of the candidate vaginal microbicide Carraguard® in South Africa | Altini | 2010 | 10.1016/j.contraception.2010.04.019 | Clinical Trial | General Population | South Africa | Southern |
| 44 | HIV prevalence and incidence in people 50 years and older in rural South Africa | Wallrauch | 2010 | NA | Prospective Cohort | General Population | South Africa | Southern |
| 45 | Combined Impact of Sexual Risk Behaviors for HIV Seroconversion Among Women in Durban, South Africa: Implications for Prevention Policy and Planning | Wand | 2011 | 10.1007/s10461-010-9845-2 | Prospective Cohort | General Population | South Africa | Southern |
| 46 | A simpler tool for estimation of HIV incidence from cross-sectional, age-specific prevalence data | Rajan | 2011 | 10.1136/jech.2009.091959 | Secondary Trial Analysis | High Risk | Nigeria | Western/Central |
| 47 | Diagnosis and counselling of patients with acute HIV infection in South Africa | Wolpaw | 2011 | 10.1136/sti.2009.041475 | Prospective Cohort | High Risk | South Africa | Southern |
| 48 | HIV-1 transmission among HIV-1 discordant couples before and after the introduction of antiretroviral therapy | Reynolds | 2011 | 10.1097/QAD.0b013e3283437c2b | Prospective Cohort | Serodiscordant Couple | Uganda | Eastern |
| 49 | High Human Immunodeficiency Virus Incidence in a Cohort of Rwandan Female Sex Workers | Braunstein | 2011 | 10.1097/OLQ.0b013e31820b8eba | Prospective Cohort | Sex Worker | Rwanda | Eastern |
| 50 | Incident HIV Infection in Pregnant and Lactating Women and Its Effect on Mother-to-Child Transmission in South Africa | Moodley | 2011 | 10.1093/infdis/jir017 | Prospective Cohort | Pregnant Women | South Africa | Southern |
| 51 | Pregnancy and HIV transmission among HIV‐discordant couples in a clinical trial in Kisumu, Kenya | Brubaker | 2011 | 10.1111/j.1468-1293.2010.00884.x | Secondary Trial Analysis | Serodiscordant Couple | Kenya | Eastern |
| 52 | Social Capital and Women's Reduced Vulnerability to HIV Infection in Rural Zimbabwe | Gregson | 2011 | 10.1111/j.1728-4457.2011.00413.x | Prospective Cohort | General Population | Zimbabwe | Southern |
| 53 | Did national HIV prevention programs contribute to HIV decline in Eastern Zimbabwe? Evidence from a prospective community survey | Gregson | 2011 | 10.1097/OLQ.0b013e3182080877 | Prospective Cohort | General Population | Zimbabwe | Southern |
| 54 | Recent HIV Type 1 Infection Among Participants in a Same-Day Mobile Testing Pilot Study in Zimbabwe | Truong | 2011 | 10.1089/aid.2010.0249 | Cross Sectional Incidence | General Population | Zimbabwe | Southern |
| 55 | High prevalent and incident HIV-1 and herpes simplex virus 2 infection among male migrant and non-migrant sugar farm workers in Zambia | Heffron | 2011 | 10.1136/sti.2010.045617 | Prospective Cohort | Other | Zambia | Southern |
| 56 | Prevalence and Incidence of HIV in a Rural Community-Based HIV Vaccine Preparedness Cohort in Masaka, Uganda | Ruzagira | 2011 | 10.1371/journal.pone.0020684 | Prospective Cohort | General Population | Uganda | Eastern |
| 57 | Sexual health, HIV risk, and retention in an adolescent HIV-prevention trial preparatory cohort | Jaspan | 2011 | 10.1016/j.jadohealth.2010.10.009 | Prospective Cohort | General Population | South Africa | Southern |
| 58 | Safety and efficacy of the HVTN 503/Phambili Study of a clade-B-based HIV-1 vaccine in South Africa: a double-blind, randomised, placebo-controlled test-of-concept phase 2b study | Gray | 2011 | 10.1016/S1473-3099(11)70098-6 | Clinical Trial | General Population | South Africa | Southern |
| 59 | Effect of concurrent sexual partnerships on rate of new HIV infections in a high-prevalence, rural South African population: a cohort study | Tanser | 2011 | 10.1016/S0140-6736(11)60779-4 | Prospective Cohort | General Population | South Africa | Southern |
| 60 | HIV Incidence and Risk Factors for Acquisition in HIV Discordant Couples in Masaka, Uganda: An HIV Vaccine Preparedness Study | Ruzagira | 2011 | 10.1371/journal.pone.0024037 | Prospective Cohort | Serodiscordant Couple | Uganda | Eastern |
| 61 | High Burden of Prevalent and Recently Acquired HIV among Female Sex Workers and Female HIV Voluntary Testing Center Clients in Kigali, Rwanda | Braunstein | 2011 | 10.1371/journal.pone.0024321 | Cross Sectional Incidence | Multiple Risk Groups | Rwanda | Eastern |
| 62 | Anal Sex, Vaginal Practices, and HIV Incidence in Female Sex Workers in Urban Kenya: Implications for the Development of Intravaginal HIV Prevention Methods | Priddy | 2011 | 10.1089/aid.2010.0362 | Prospective Cohort | Sex Worker | Kenya | Eastern |
| 63 | Incidence of HIV in Windhoek, Namibia: Demographic and Socio-Economic Associations | Aulagnier | 2011 | 10.1371/journal.pone.0025860 | Serial Crosssectional Survey | General Population | Namibia | Southern |
| 64 | The relative contribution of viral and bacterial sexually transmitted infections on HIV acquisition in southern African women in the Methods for Improving Reproductive Health in Africa study | Venkatesh | 2011 | 10.1258/ijsa.2010.010385 | Nested Case Control | General Population | Multiple Countries | Multiple Countries |
| 65 | Risk factors for HIV-1 infection in a longitudinal, prospective cohort of adults from the Mbeya Region, Tanzania | Geis | 2011 | 10.1097/QAI.0b013e3182118fa3 | Prospective Cohort | General Population | Tanzania | Eastern |
| 66 | Dual Testing Algorithm of BED-CEIA and AxSYM Avidity Index Assays Performs Best in Identifying Recent HIV Infection in a Sample of Rwandan Sex Workers | Braunstein | 2011 | 10.1371/journal.pone.0018402 | Cross Sectional Incidence/Prospective Cohort | Sex Worker | Rwanda | Eastern |
| 67 | Safety and effectiveness of BufferGel and 0.5% PRO2000 gel for the prevention of HIV infection in women | Karim | 2011 | 10.1097/QAD.0b013e32834541d9 | Clinical Trial | General Population | South Africa, Malawi, Multiple Countries | Multiple Countries |
| 68 | Estimates of human immunodeficiency virus incidence among female sex workers in north central Nigeria: implications for HIV clinical trials | Forbi | 2011 | 10.1016/j.trstmh.2011.08.001 | Cross Sectional Incidence | Sex Worker | Nigeria | Western/Central |
| 69 | Stabilizing HIV prevalence masks high HIV incidence rates amongst rural and urban women in KwaZulu-Natal, South Africa | Karim | 2011 | 10.1093/ije/dyq176 | Prospective Cohort | General Population | South Africa | Southern |
| 70 | Improved detection of incident HIV infection and uptake of PMTCT services in labor and delivery in a high HIV prevalence setting | Kieffer | 2011 | 10.1097/QAI.0b013e31821acc6e | Clinical Trial | Pregnant Women | eSwatini | Southern |
| 71 | Prevention of HIV-1 Infection with Early Antiretroviral Therapy | Cohen | 2011 | 10.1056/NEJMoa1105243 | Clinical Trial | Serodiscordant Couple | Kenya, Zimbabwe, Malawi, Botswana, South Africa | Eastern/Southern |
| 72 | HIV Prevalence and Incidence among Sexually Active Females in Two Districts of South Africa to Determine Microbicide Trial Feasibility | Nel | 2011 | 10.1371/journal.pone.0021528 | Prospective Cohort/Cross Sectional Incidence | General Population | South Africa | Southern |
| 73 | Changes in sexual risk behavior before and after HIV seroconversion in Southern African women enrolled in a HIV prevention trial | Venkatesh | 2011 | 10.1097/QAI.0b013e318220379b | Secondary Trial Analysis | General Population | South Africa, Zimbabwe | Multiple Countries |
| 74 | The effectiveness of male circumcision for HIV prevention and effects on risk behaviors in a posttrial follow-up study | Gray | 2012 | 10.1097/QAD.0b013e3283504a3f. | Prospective Cohort/Secondary Trial Analysis | General Population | Uganda | Eastern |
| 75 | Prevalence of seroconversion symptoms and relationship to set-point viral load: findings from a subtype C epidemic, 1995–2009 | Sullivan | 2012 | 10.1097/QAD.0b013e32834ed8c8 | Prospective Cohort | Serodiscordant Couple | Zambia | Southern |
| 76 | Identifying at-risk populations in Kenya and South Africa: HIV incidence in cohorts of men who report sex with men, sex workers, and youth | Price | 2012 | 10.1097/QAI.0b013e31823d8693 | Prospective Cohort | Multiple Risk Groups | South Africa, Kenya | Eastern/Southern |
| 77 | Characterization of Acute HIV-1 Infection in High-Risk Nigerian Populations | Charurat | 2012 | 10.1093/infdis/jis103 | Prospective Cohort | General Population | Nigeria | Western/Central |
| 78 | HIV Incidence Remains High in KwaZulu-Natal, South Africa: Evidence from Three Districts | Nel | 2012 | 10.1371/journal.pone.0035278 | Prospective Cohort/Cross Sectional Incidence | General Population | South Africa | Southern |
| 79 | Circumcision status and incident herpes simplex virus type 2 infection, genital ulcer disease, and HIV infection | Mehta | 2012 | 10.1097/QAD.0b013e328352d116 | Secondary Trial Analysis | General Population | Kenya | Eastern |
| 80 | High HIV incidence and socio-behavioral risk patterns in fishing communities on the shores of Lake Victoria, Uganda | Seeley | 2012 | 10.1097/OLQ.0b013e318251555d | Prospective Cohort | Fisher Folk | Uganda | Eastern |
| 81 | High HIV Incidence and Sexual Behavior Change among Pregnant Women in Lilongwe, Malawi: Implications for the Risk of HIV Acquisition | Keating | 2012 | 10.1371/journal.pone.0039109 | Retrospective Cohort | Pregnant Women | Malawi | Southern |
| 82 | HIV Incidence Among Non-Pregnant Women Living in Selected Rural, Semi-Rural and Urban Areas in Kwazulu-Natal, South Africa | Ramjee | 2012 | 10.1007/s10461-011-0043-7 | Secondary Trial Analysis | General Population | South Africa | Southern |
| 83 | HIV Incidence in Young Girls in KwaZulu-Natal, South Africa-Public Health Imperative for Their Inclusion in HIV Biomedical Intervention Trials | Karim | 2012 | 10.1007/s10461-012-0209-y | Prospective Cohort | General Population | South Africa | Southern |
| 84 | Periodical antibiotic treatment for the control of gonococcal and chlamydial infections among sex workers in Benin and Ghana: a cluster-randomized placebo-controlled trial | Labbe | 2012 | 10.1097/OLQ.0b013e318244aaa0 | Clinical Trial | Sex Worker | Multiple Countries | Multiple Countries |
| 85 | Symptomatic Vaginal Discharge Is a Poor Predictor of Sexually Transmitted Infections and Genital Tract Inflammation in High-Risk Women in South Africa | Mlisana | 2012 | 10.1093/infdis/jis298 | Prospective Cohort | High Risk | South Africa | Southern |
| 86 | The Rates of HIV Superinfection and Primary HIV Incidence in a General Population in Rakai, Uganda | Redd | 2012 | 10.1093/infdis/jis325 | Prospective Cohort | General Population | Uganda | Eastern |
| 87 | Trends in the uptake of voluntary counselling and testing for HIV in rural Tanzania in the context of the scale up of antiretroviral therapy | Isingo | 2012 | 10.1111/j.1365-3156.2011.02877.x | Serial Crosssectional Survey | General Population | Tanzania | Eastern |
| 88 | Antiretroviral Preexposure Prophylaxis for Heterosexual HIV Transmission in Botswana | Thigpen | 2012 | 10.1056/NEJMoa1110711 | Clinical Trial | General Population | Botswana | Southern |
| 89 | Antiretroviral Prophylaxis for HIV Prevention in Heterosexual Men and Women | Baeten | 2012 | 10.1056/NEJMoa1108524 | Clinical Trial | Serodiscordant Couple | Kenya, Uganda | Multiple Countries |
| 90 | Preexposure Prophylaxis for HIV Infection among African Women | Van Damme | 2012 | 10.1056/NEJMoa1202614 | Clinical Trial | High Risk | South Africa, Tanzania, Kenya | Eastern/Southern |
| 91 | Declining Rate of Infection with Maternal Human Immunodeficiency Virus at DeliveryUnits in North-Central Nigeria | Imade | 2013 | NA | Prospective Cohort | Pregnant Women | Nigeria | Western/Central |
| 92 | High HIV-1 incidence, correlates of HIV-1 acquisition, and high viral loads following seroconversion among MSM | Sanders | 2013 | 10.1097/QAD.0b013e32835b0f81 | Prospective Cohort | MSM | Kenya | Eastern |
| 93 | HIV postexposure prophylaxis in an urban population of female sex workers in Nairobi, Kenya | Izulla | 2013 | 10.1097/QAI.0b013e318278ba1b | Prospective Cohort | Sex Worker | Kenya | Eastern |
| 94 | HIV-1 Transmission within Marriage in Rural Uganda: A Longitudinal Study | Biraro | 2013 | 10.1371/journal.pone.0055060 | Prospective Cohort | Serodiscordant Couple | Uganda | Eastern |
| 95 | Oral and injectable contraception use and risk of HIV acquisition among women in sub-Saharan Africa | McCoy | 2013 | 10.1097/QAD.0b013e32835da401 | Secondary Trial Analysis | General Population | Multiple Countries | Multiple Countries |
| 96 | Alcohol use, mycoplasma genitalium, and other STIs associated With HIV incidence among women at high risk in Kampala, Uganda | Vanderpitte | 2013 | 10.1097/QAI.0b013e3182777167 | Prospective Cohort | Sex Worker* | Uganda | Eastern |
| 97 | Short Communication: HIV Type 1 Transmitted Drug Resistance and Evidence of Transmission Clusters Among Recently Infected Antiretroviral-Naive Individuals from Ugandan Fishing Communities of Lake Victoria | Nazziwa | 2013 | 10.1089/aid.2012.0123 | Prospective Cohort | Fisher Folk | Uganda | Eastern |
| 98 | Evidence for a contribution of the community response to HIV decline in eastern Zimbabwe? | Gregson | 2013 | 10.1080/09540121.2012.748171 | Prospective Cohort | General Population | Zimbabwe | Southern |
| 99 | HIV Prevalence and Incidence Among Women at Higher Risk of Infection in Addis Ababa, Ethiopia | Combes | 2013 | 10.1089/aid.2012.0163 | Cross Sectional Incidence | General Population | Ethiopia | Eastern |
| 100 | Behavioral, Biological, and Demographic Risk and Protective Factors for New HIV Infections Among Youth in Rakai, Uganda | Santelli | 2013 | 10.1097/QAI.0b013e3182926795 | Prospective Cohort | General Population | Uganda | Eastern |
| 101 | Estimation of HIV Incidence in a Large, Community-Based, Randomized Clinical Trial: NIMH Project Accept (HIV Prevention Trials Network 043) | Laeyendecker | 2013 | 10.1371/journal.pone.0068349 | Cross Sectional Incidence | General Population | South Africa, Tanzania, Zimbabwe | Multiple Countries |
| 102 | The Epidemiology of HIV and HSV-2 Infections among Women Participating in Microbicide and Vaccine Feasibility Studies in Northern Tanzania | Kapiga | 2013 | 10.1371/journal.pone.0068825 | Prospective Cohort | High Risk | Tanzania | Eastern |
| 103 | Long-term consistent use of a vaginal microbicide gel among HIV-1 sero-discordant couples in a phase III clinical trial (MDP 301) in rural south-west Uganda | Abaasa | 2013 | 10.1186/1745-6215-14-33 | Clinical Trial | Serodiscordant Couple | Uganda | Eastern |
| 104 | Association of the ANRS-12126 Male Circumcision Project with HIV Levels among Men in a South African Township: Evaluation of Effectiveness using Cross-sectional Surveys | Auvert | 2013 | 10.1371/journal.pmed.1001509 | Cross Sectional Incidence | General Population | South Africa | Southern |
| 105 | Adherence to Antiretroviral Prophylaxis for HIV Prevention: A Substudy Cohort within a Clinical Trial of Serodiscordant Couples in East Africa | Haberer | 2013 | 10.1371/journal.pmed.1001511 | Prospective Cohort | Serodiscordant Couple | Uganda | Eastern |
| 106 | Non-consensual Sex and Association with Incident HIV Infection Among Women: A Cohort Study in Rural Uganda, 1990–2008 | Birdthistle | 2013 | 10.1007/s10461-012-0378-8 | Prospective Cohort | General Population | Uganda | Eastern |
| 107 | Effects of hormonal contraceptive use on HIV acquisition and transmission among HIV-discordant couples | Lutalo | 2013 | 10.1097/QAD.0000000000000045 | Prospective Cohort | Serodiscordant Couple | Uganda | Eastern |
| 108 | Effect of HSV‐2 on population‐level trends in HIV incidence in Uganda between 1990 and 2007 | Biraro | 2013 | 10.1111/tmi.12176 | Prospective Cohort | General Population | Uganda | Eastern |
| 109 | Declining HIV-1 prevalence and incidence among Police Officers – a potential cohort for HIV vaccine trials, in Dar es Salaam, Tanzania | Munseri | 2013 | 10.1186/1471-2458-13-722 | Prospective Cohort | Other | Tanzania | Eastern |
| 110 | Efficacy of preexposure prophylaxis for HIV-1 prevention among high-risk heterosexuals: subgroup analyses from a randomized trial | Murnane | 2013 | 10.1097/QAD.0b013e3283629037 | Secondary Trial Analysis | Serodiscordant Couple | Multiple Countries | Multiple Countries |
| 111 | Assessing the effect of HIV counselling and testing on HIV acquisition among South African youth | Rosenberg | 2013 | 10.1097/01.aids.0000432454.68357.6a | Prospective Cohort | General Population | South Africa | Southern |
| 112 | Evaluating HIV prevention efforts using semiparametric regression models: results from a large cohort of women participating in an HIV prevention trial from KwaZulu‐Natal, South Africa | Wand | 2013 | 10.7448/IAS.16.1.18589 | Secondary Trial Analysis | General Population | South Africa | Southern |
| 113 | The long-term efficacy of medical male circumcision against HIV acquisition | Mehta | 2013 | 10.1097/01.aids.0000432444.30308.2d | Prospective Cohort | General Population | Kenya | Eastern |
| 114 | Prevalence, Incidence and Determinants of Herpes Simplex Virus Type 2 Infection among HIV-Seronegative Women at High-Risk of HIV Infection: A Prospective Study in Beira, Mozambique | Meque | 2014 | 10.1371/journal.pone.0089705 | Prospective Cohort | High Risk | Mozambique | Southern |
| 115 | Effects of hazardous and harmful alcohol use on HIV incidence and sexual behaviour: a cohort study of Kenyan female sex workers | Chersich | 2014 | 10.1186/1744-8603-10-22 | Prospective Cohort | Sex Worker | Kenya | Eastern |
| 116 | Acceptability and feasibility of serial HIV antibody testing during pregnancy/postpartum and male partner testing in Tororo, Uganda | Kim | 2014 | 10.1080/09540121.2013.824536 | Prospective Cohort | Pregnant Women | Uganda | Eastern |
| 117 | High HIV incidence in the postpartum period sustains vertical transmission in settings with generalized epidemics: a cohort study in Southern Mozambique | De Schacht | 2014 | 10.7448/IAS.17.1.18808 | Prospective Cohort | Pregnant Women | Mozambique | Southern |
| 118 | Risk of HIV acquisition among circumcised and uncircumcised young men with penile human papillomavirus infection | Rositch | 2014 | 10.1097/QAD.0000000000000092 | Prospective Cohort | General Population | Kenya | Eastern |
| 119 | Early adolescent pregnancy increases risk of incident HIV infection in the Eastern Cape, South Africa: a longitudinal study | Christofides | 2014 | 10.7448/IAS.17.1.18585 | Secondary Trial Analysis | General Population | South Africa | Southern |
| 120 | The impact of voluntary counselling and testing services on sexual behaviour change and HIV incidence: observations from a cohort study in rural Tanzania | Cawley | 2014 | 10.1186/1471-2334-14-159 | Serial Crosssectional Survey | General Population | Tanzania | Eastern |
| 121 | Incidence of TB and HIV in Prospectively Followed Household Contacts of TB Index Patients in South Africa | Van Schalkwyk | 2014 | 10.1371/journal.pone.0095372 | Prospective Cohort | Other | South Africa | Southern |
| 122 | HIV Incidence and Factors Associated with Seroconversion in a Rural Community Home Based Counseling and Testing Program in Eastern Uganda | Okiria | 2014 | 10.1007/s10461-013-0502-4 | Serial Crosssectional Survey | General Population | Uganda | Eastern |
| 123 | Do age-disparate relationships drive HIV incidence in young women? Evidence from a population cohort in rural KwaZulu-Natal, South Africa | Harling | 2014 | 10.1097/QAI.0000000000000198 | Prospective Cohort | General Population | South Africa | Southern |
| 124 | Use of antiretroviral therapy in households and risk of HIV acquisition in rural KwaZulu-Natal, South Africa, 2004–12: a prospective cohort study | Vandormael | 2014 | 10.1016/S2214-109X(14)70018-X | Prospective Cohort | General Population | South Africa | Southern |
| 125 | High HIV risk in a cohort of male sex workers from Nairobi, Kenya | McKinnon | 2014 | 10.1136/sextrans-2013-051310 | Prospective Cohort | Multiple Risk Groups | Kenya | Eastern |
| 126 | Effect of community-based voluntary counselling and testing on HIV incidence and social and behavioural outcomes (NIMH Project Accept; HPTN 043): a cluster-randomised trial | Coates | 2014 | 10.1016/S2214-109X(14)70032-4 | Cross Sectional Incidence | General Population | Zimbabwe, Multiple Countries | Multiple Countries |
| 127 | Single-agent tenofovir versus combination emtricitabine plus tenofovir for pre-exposure prophylaxis for HIV-1 acquisition: an update of data from a randomised, double-blind, phase 3 trial | Baeten | 2014 | 10.1016/S1473-3099(14)70937-5 | Clinical Trial | Serodiscordant Couple | Kenya, Uganda | Multiple Countries |
| 128 | Marriage and the risk of incident HIV infection in Rakai, Uganda | Nalugoda | 2014 | 10.1097/QAI.0b013e3182a7f08a | Prospective Cohort | General Population | Uganda | Eastern |
| 129 | HIV Prevalence and Incidence in a Cohort of Women at Higher Risk for HIV Acquisition in Chókwè, Southern Mozambique | Feldblum | 2014 | 10.1371/journal.pone.0097547 | Prospective Cohort | High Risk | Mozambique | Southern |
| 130 | High Incidence of HIV-1 Infection in a General Population of Fishing Communities around Lake Victoria, Uganda | Kiwanuka | 2014 | 10.1371/journal.pone.0094932 | Prospective Cohort | Fisher Folk | Uganda | Eastern |
| 131 | An assessment of fishing communities around Lake Victoria, Uganda, as potential populations for future HIV vaccine efficacy studies: an observational cohort study | Kiwanuka | 2014 | 10.1186/1471-2458-14-986 | Prospective Cohort | Fisher Folk | Uganda | Eastern |
| 132 | Recombinant adenovirus type 5 HIV gag/pol/nef vaccine in South Africa: unblinded, long-term follow-up of the phase 2b HVTN 503/Phambili study | Gray | 2014 | 10.1016/S1473-3099(14)70020-9 | Clinical Trial | General Population | South Africa | Southern |
| 133 | Long-term follow-up of study participants from prophylactic HIV vaccine clinical trials in Africa | Schmidt | 2014 | 10.4161/hv.27559 | Prospective Cohort | General Population | Multiple Countries | Multiple Countries |
| 134 | Rates of HIV-1 superinfection and primary HIV-1 infection are similar in female sex workers in Uganda | Redd | 2014 | 10.1097/QAD.0000000000000365 | Prospective Cohort | Sex Worker | Uganda | Eastern |
| 135 | HIV Incidence in a Cohort of Women at Higher Risk in Beira, Mozambique: Prospective Study 2009–2012 | Dube | 2014 | 10.1371/journal.pone.0084979 | Prospective Cohort | High Risk | Mozambique | Southern |
| 136 | High Rates of HIV Seroconversion in Pregnant Women and Low Reported Levels of HIV Testing among Male Partners in Southern Mozambique: Results from a Mixed Methods Study | DeSchacht | 2014 | 10.1371/journal.pone.0115014 | Prospective Cohort | Pregnant Women | Mozambique | Southern |
| 137 | Factors associated with incident HIV infection versus prevalent infection among youth in Rakai, Uganda | Edelstein | 2015 | 10.1016/j.jegh.2014.09.003 | Prospective Cohort | General Population | Uganda | Eastern |
| 138 | Spatial clustering of “measured” and “unmeasured” risk factors for HIV infections in hyper-endemic communities in KwaZulu-Natal, South Africa: results from geoadditive models | Wand | 2015 | 10.1080/09540121.2015.1096896 | Secondary Trial Analysis | General Population | South Africa | Southern |
| 139 | Creating an African HIV Clinical Research and Prevention Trials Network: HIV Prevalence, Incidence and Transmission | Kamali | 2015 | 10.1371/journal.pone.0116100 | Prospective Cohort | Multiple Risk Groups | South Africa, Uganda | Eastern/Southern |
| 140 | Sero-conversion rate of Syphilis and HIV among pregnant women attending antenatal clinic in Tanzania: a need for re-screening at delivery | Lawi | 2015 | 10.1186/s12884-015-0434-2 | Prospective Cohort | Pregnant Women | Tanzania | Eastern |
| 141 | Tenofovir-Based Preexposure Prophylaxis for HIV Infection among African Women | Marrazzo | 2015 | 10.1056/NEJMoa1402269 | Clinical Trial | General Population | South Africa, Multiple Countries | Multiple Countries |
| 142 | Incidence of HIV and the Prevalence of HIV, Hepatitis B and Syphilis among Youths in Maputo, Mozambique: A Cohort Study | Viegas | 2015 | 10.1371/journal.pone.0121452 | Prospective Cohort | General Population | Mozambique | Southern |
| 143 | Biological impact of recurrent sexually transmitted infections on HIV seroconversion among women in South Africa: results from frailty models | Wand | 2015 | 10.7448/IAS.18.1.19866 | Secondary Trial Analysis | General Population | South Africa | Southern |
| 144 | Hormonal contraception does not increase women’s HIV acquisition risk in Zambian discordant couples, 1994–2012 | Wall | 2015 | 10.1016/j.contraception.2015.02.004 | Prospective Cohort | Serodiscordant Couple | Zambia | Southern |
| 145 | Trends in HIV acquisition, risk factors and prevention policies among youth in Uganda, 1999-2011 | Santelli | 2015 | 10.1097/QAD.0000000000000533 | Prospective Cohort | General Population | Uganda | Eastern |
| 146 | Effectiveness of an integrated intimate partner violence and HIV prevention intervention in Rakai, Uganda: analysis of an intervention in an existing cluster randomised cohort | Wagman | 2015 | 10.1016/S2214-109X(14)70344-4 | Clinical Trial | General Population | Uganda | Eastern |
| 147 | Impact of Maternal HIV Seroconversion during Pregnancy on Early Mother to Child Transmission of HIV (MTCT) Measured at 4-8 Weeks Postpartum in South Africa 2011-2012: A National Population-Based Evaluation | Dinh | 2015 | 10.1371/journal.pone.0125525 | Prospective Cohort | Pregnant Women | South Africa | Southern |
| 148 | Risk of HIV-1 acquisition among women who use different types of injectable progestin contraception in South Africa: a prospective cohort study | Noguchi | 2015 | 10.1016/S2352-3018(15)00058-2 | Secondary Trial Analysis | General Population | South Africa | Southern |
| 149 | HIV acquisition during pregnancy and postpartum is associated with genital infections and partnership characteristics | Kinuthia | 2015 | 10.1097/QAD.0000000000000793 | Prospective Cohort | Pregnant Women | Kenya | Eastern |
| 150 | Age-Disparate Partnerships and Risk of HIV-1 Acquisition Among South African Women Participating in the VOICE Trial | Balkus | 2015 | 10.1097/QAI.0000000000000715 | Secondary Trial Analysis | General Population | South Africa | Southern |
| 151 | A 15-year study of the impact of community antiretroviral therapy coverage on HIV incidence in Kenyan female sex workers | McClelland | 2015 | 10.1097/QAD.0000000000000829 | Prospective Cohort | Sex Worker | Kenya | Eastern |
| 152 | Lack of Effectiveness of Antiretroviral Therapy in Preventing HIV Infection in Serodiscordant Couples in Uganda: An Observational Study | Birungi | 2015 | 10.1371/journal.pone.0132182 | Prospective Cohort | Serodiscordant Couple | Uganda | Eastern |
| 153 | Partner Age-Disparity and HIV Incidence Risk for Older Women in Rural South Africa | Harling | 2015 | 10.1007/s10461-014-0952-3 | Prospective Cohort | General Population | South Africa | Southern |
| 154 | Risk Factors for HIV Acquisition in High Risk Women in a Generalised Epidemic Setting | Naicker | 2015 | 10.1007/s10461-015-1002-5 | Prospective Cohort | Sex Worker | South Africa | Southern |
| 155 | Cascade of HIV care and population viral suppression in a high-burden region of Kenya | Maman | 2015 | 10.1097/QAD.0000000000000741 | Cross Sectional Incidence | General Population | Kenya | Eastern |
| 156 | Re-Testing and Seroconversion Among HIV Testing and Counseling Clients in Lesotho | Grabbe | 2015 | 10.1521/aeap.2015.27.4.350 | Retrospective Cohort | General Population | Lesotho | Southern |
| 157 | High Transmitter CD4+ T-Cell Count Shortly after the Time of Transmission in a Study of African Serodiscordant Couples | Karita | 2015 | 10.1371/journal.pone.0134438 | Prospective Cohort | Serodiscordant Couple | Rwanda, Uganda, Zambia | Multiple Countries |
| 158 | The Prevalence of Human Immunodeficiency Virus Infection among Pregnant Women inLabour with Unknown Status and those with Negative Status Early in the Index Pregnancyin a Tertiary Hospital in Nigeria | Ukaire | 2015 | N/A | Prospective Cohort | Pregnant Women | Nigeria | Western/Central |
| 159 | Continued Follow-Up of Phambili Phase 2b Randomized HIV-1 Vaccine Trial Participants Supports Increased HIV-1 Acquisition among Vaccinated Men | Moodie | 2015 | 10.1371/journal.pone.0137666 | Clinical Trial/Prospective Cohort | General Population | South Africa | Southern |
| 160 | HIV prevention and care services for female sex workers: efficacy of a targeted community-based intervention in Burkina Faso | Traore | 2015 | 10.7448/IAS.18.1.20088 | Prospective Cohort | Sex Worker | Burkina Faso | Western/Central |
| 161 | Factors associated with dropout in a long term observational cohort of fishing communities around lake Victoria, Uganda | Abaasa | 2015 | 10.1186/S13104-015-1804-6 | Prospective Cohort | Fisher Folk | Uganda | Eastern |
| 162 | Investigation of HIV Incidence Rates in a High-Risk, High-Prevalence Kenyan Population: Potential Lessons for Intervention Trials and Programmatic Strategies | Mdodo | 2016 | 10.1177/2325957413511667 | Prospective Cohort | Multiple Risk Groups | Kenya | Eastern |
| 163 | Estimating HIV Incidence during Pregnancy and Knowledge of Prevention of Mother-to-Child Transmission with an Ad Hoc Analysis of Potential Cofactors | Egbe | 2016 | 10.1155/2016/7397695 | Prospective Cohort | Pregnant Women | Cameroon | Western/Central |
| 164 | Closer to 90-90-90. The cascade of care after 10 years of ART scale-up in rural Malawi: a population study | Maman | 2016 | 10.7448/IAS.19.1.20673 | Cross Sectional Incidence | General Population | Malawi | Southern |
| 165 | New insights into HIV epidemic in South Africa: key findings from the National HIV Prevalence, Incidence and Behaviour Survey, 2012 | Zuma | 2016 | 10.2989/16085906.2016.1153491 | Cross Sectional Incidence | General Population | South Africa | Southern |
| 166 | Association between injectable progestin-only contraceptives and HIV acquisition and HIV target cell frequency in the female genital tract in South African women: a prospective cohort study | Byrne | 2016 | 10.1016/S1473-3099(15)00429-6 | Prospective Cohort | General Population | South Africa | Southern |
| 167 | Comparison of HIV incidence estimated in clinical trial and observational cohort settings in a high risk fishing population in Uganda: Implications for sample size estimates | Abaasa | 2016 | 10.1016/j.vaccine.2016.02.048 | Prospective Cohort | Fisher Folk | Uganda | Eastern |
| 168 | Implementation and Operational Research: Cohort Analysis of Program Data to Estimate HIV Incidence and Uptake of HIV-Related Services Among Female Sex Workers in Zimbabwe, 2009-2014 | Hargreaves | 2016 | 10.1097/QAI.0000000000000920 | Prospective Cohort | Sex Worker | Zimbabwe | Southern |
| 169 | Prospective Study of Acute HIV-1 Infection in Adults in East Africa and Thailand | Robb | 2016 | 10.1056/NEJMoa1508952 | Prospective Cohort | Multiple Risk Groups | Kenya, Tanzania, Uganda, Multiple Countries | Multiple Countries |
| 170 | Postcoital penile washing and the risk of HIV acquisition in uncircumcised men | Makumbi | 2016 | 10.1097/QAD.0000000000001097 | Prospective Cohort | Other | Uganda | Eastern |
| 171 | Development of a prospective cohort of HIV Exposed Sero-Negative (HESN) individuals in Jos Nigeria | Osawe | 2016 | 10.1186/s12879-016-1649-1 | Prospective Cohort | Serodiscordant Couple | Nigeria | Western/Central |
| 172 | Integrated Delivery of Antiretroviral Treatment and Pre-exposure Prophylaxis to HIV-1–Serodiscordant Couples: A Prospective Implementation Study in Kenya and Uganda | Baeten | 2016 | 10.1371/journal.pmed.1002099 | Prospective Cohort | Serodiscordant Couple | Multiple Countries | Multiple Countries |
| 173 | HIV Transmission Risk Persists During the First 6 Months of Antiretroviral Therapy | Mujugira | 2016 | 10.1097/QAI.0000000000001019 | Secondary Trial Analysis | Serodiscordant Couple | Multiple Countries | Multiple Countries |
| 174 | Antiretroviral Therapy to Prevent HIV Acquisition in Serodiscordant Couples in a Hyperendemic Community in Rural South Africa | Oldenburg | 2016 | 10.1093/cid/ciw335 | Prospective Cohort | Serodiscordant Couple | South Africa | Southern |
| 175 | Heterogeneity of HIV incidence: a comparative analysis between fishing communities and in a neighbouring rural general population, Uganda, and implications for HIV control | Kamali | 2016 | 10.1136/sextrans-2015-052179 | Prospective Cohort | Multiple Risk Groups | Uganda | Eastern |
| 176 | Hormonal Contraceptive Use Among HIV-Positive Women and HIV Transmission Risk to Male Partners, Zambia, 1994–2012 | Wall | 2016 | 10.1093/infdis/jiw322 | Prospective Cohort | Serodiscordant Couple | Zambia | Southern |
| 177 | Association of Medical Male Circumcision and Antiretroviral Therapy Scale-up With Community HIV Incidence in Rakai, Uganda | Kong | 2016 | 10.1001/jama.2016.7292 | Prospective Cohort | General Population | Uganda | Eastern |
| 178 | Detection of Acute and Early HIV-1 Infections in an HIV Hyper-Endemic Area with Limited Resources | Mayaphi | 2016 | 10.1371/journal.pone.0164943 | Cross Sectional Incidence | Multiple Risk Groups | South Africa | Southern |
| 179 | Effect of Wuchereria bancrofti infection on HIV incidence in southwest Tanzania: a prospective cohort study | Kroidl | 2016 | 10.1016/S0140-6736(16)31252-1 | Prospective Cohort | General Population | Tanzania | Eastern |
| 180 | Rising Levels of HIV Infection in Older Adults in Eastern Zimbabwe | Negin | 2016 | 10.1371/journal.pone.0162967 | Serial Crosssectional Survey | General Population | Zimbabwe | Southern |
| 181 | The effect of a conditional cash transfer on HIV incidence in young women in rural South Africa (HPTN 068): a phase 3, randomised controlled trial | Pettifor | 2016 | 10.1016/S2214-109X(16)30253-4 | Clinical Trial | General Population | South Africa | Southern |
| 182 | Use of a Vaginal Ring Containing Dapivirine for HIV-1 Prevention in Women | Baeten | 2016 | 10.1056/NEJMoa1506110 | Clinical Trial | General Population | South Africa, Multiple Countries | Multiple Countries |
| 183 | Safety and Efficacy of a Dapivirine Vaginal Ring for HIV Prevention in Women | Nel | 2016 | 10.1056/NEJMoa1602046 | Clinical Trial | General Population | Multiple Countries | Multiple Countries |
| 184 | Prevalence of Herpes Simplex Virus 2 (HSV-2) infection and associated risk factors in a cohort of HIV negative women in Durban, South Africa | Daniels | 2016 | 10.1186/s13104-016-2319-5 | Secondary Trial Analysis | General Population | South Africa | Southern |
| 185 | Brief Report: High HIV Incidence in a South African Community of Men Who Have Sex With Men: Results From the Mpumalanga Men's Study, 2012-2015. | Lane | 2016 | 10.1097/QAI.0000000000001162 | Serial Crosssectional Survey | MSM | South Africa | Southern |
| 186 | The prevalence and determinants of HIV seroconversion among booked ante natal clients in the University of Uyo teaching hospital, Uyo Akwa Ibom State, Nigeria | Nyoyoko | 2016 | 10.11604/pamj.2016.25.247.6715 | Prospective Cohort | Pregnant Women | Nigeria | Western/Central |
| 187 | Alignment of adherence and risk for HIV acquisition in a demonstration project of pre‐exposure prophylaxis among HIV serodiscordant couples in Kenya and Uganda: a prospective analysis of prevention‐effective adherence | Haberer | 2016 | 10.7448/IAS.20.1.21842 | Prospective Cohort | Serodiscordant Couple | Multiple Countries | Multiple Countries |
| 188 | Space-time migration patterns and risk of HIV acquisition in rural South Africa | Dobra | 2017 | 10.1097/QAD.0000000000001292 | Prospective Cohort | General Population | South Africa | Southern |
| 189 | Identifying Factors Associated with Low-Adherence and Subsequent HIV Seroconversions Among South African Women Enrolled in a Biomedical Intervention Trial | Wand | 2017 | 10.1007/s10461-016-1471-1 | Secondary Trial Analysis | General Population | South Africa | Southern |
| 190 | Swaziland HIV Incidence Measurement Survey (SHIMS): a prospective national cohort study | Justman | 2017 | 10.1016/S2352-3018(16)30190-4 | Prospective Cohort | General Population | eSwatini | Southern |
| 191 | Population attributable fraction of incident HIV infections associated with alcohol consumption in fishing communities around Lake Victoria, Uganda | Kiwanuka | 2017 | 10.1371/journal.pone.0171200 | Prospective Cohort | Fisher Folk | Uganda | Eastern |
| 192 | High HIV prevalence and incidence among women in Southern Mozambique: Evidence from the MDP microbicide feasibility study | Mocumbi | 2017 | 10.1371/journal.pone.0173243 | Prospective Cohort | General Population | Mozambique | Southern |
| 193 | A Risk Assessment Tool for Identifying Pregnant and Postpartum Women Who May Benefit From Preexposure Prophylaxis | Pintyre | 2017 | 10.1093/cid/ciw850 | Prospective Cohort | Pregnant Women | Kenya | Eastern |
| 194 | Estimating HIV Incidence Using a Cross-Sectional Survey: Comparison of Three Approaches in a Hyperendemic Setting, Ndhiwa Subcounty, Kenya, 2012 | Blaizot | 2017 | 10.1089/aid.2016.0123 | Cross Sectional Incidence | General Population | Kenya | Eastern |
| 195 | Incident HIV during pregnancy and early postpartum period: a population-based cohort study in a rural area in KwaZulu-Natal, South Africa | Chetty | 2017 | 10.1186/s12884-017-1421-6 | Retrospective Cohort | Pregnant Women | South Africa | Southern |
| 196 | Sexual partnership age pairings and risk of HIV acquisition in rural South Africa | Akullian | 2017 | 10.1097/QAD.0000000000001553 | Prospective Cohort | General Population | South Africa | Southern |
| 197 | HIV Prevention Efforts and Incidence of HIV in Uganda | Grabowski | 2017 | 10.1056/NEJMoa1702150 | Prospective Cohort | General Population | Uganda | Eastern |
| 198 | Age-disparate relationships and HIV incidence in adolescent girls and young women evidence from Zimbabwe | Schaefer | 2017 | 10.1097/QAD.0000000000001506 | Prospective Cohort | General Population | Zimbabwe | Southern |
| 199 | Is PrEP Needed for MSM in West Africa? HIV Incidence in a Prospective Multicountry Cohort | Couderc | 2017 | 10.1097/QAI.0000000000001288 | Prospective Cohort | MSM | Mali, Cote d'Ivoire, Senegal | Multiple Countries |
| 200 | Identification of acute HIV-1 infection by Hologic Aptima HIV-1 RNA Qualitative Assay | Manak | 2017 | 10.1128/JCM.00431-17 | Prospective Cohort | High Risk | Multiple Countries | Multiple Countries |
| 201 | Low HIV incidence in pregnant and postpartum women receiving a community-based combination HIV prevention intervention in a high HIV incidence setting in South Africa | Fatti | 2017 | 10.1371/journal.pone.0181691 | Prospective Cohort | Pregnant Women | South Africa | Southern |
| 202 | The effect of school attendance and school dropout on incident HIV and HSV-2 among young women in rural South Africa enrolled in HPTN 068 | Stoner | 2017 | 10.1097/QAD.0000000000001584 | Secondary Trial Analysis | General Population | South Africa | Southern |
| 203 | A comparison of self-report and antiretroviral detection to inform estimates of antiretroviral therapy coverage, viral load suppression and HIV incidence in Kwazulu-Natal, South Africa | Huerga | 2017 | 10.1186/s12879-017-2740-y | Cross Sectional Incidence | General Population | South Africa | Southern |
| 204 | Documenting and explaining the HIV decline in east Zimbabwe: the Manicaland General Population Cohort | Gregson | 2017 | 10.1136/bmjopen-2017-015898 | Prospective Cohort | General Population | Zimbabwe | Southern |
| 205 | HIV Incidence and Predictors of HIV Acquisition From an Outside Partner in Serodiscordant Couples in Lusaka, Zambia | Joseph Davey | 2017 | 10.1097/QAI.0000000000001494 | Prospective Cohort | Serodiscordant Couple | Zambia | Southern |
| 206 | IP-10 Levels as an Accurate Screening Tool to Detect Acute HIV Infection in Resource-Limited Settings | Pastor | 2017 | 10.1038/s41598-017-08218-0 | Prospective Cohort | General Population | Mozambique | Southern |
| 207 | Difficult decisions: Evaluating individual and couple-level fertility intentions and HIV acquisition among HIV serodiscordant couples in Zambia | Joseph Davey | 2018 | 10.1371/journal.pone/0189869 | Prospective Cohort | Serodiscordant Couple | Zambia | Southern |
| 208 | Brief Report: PrEP Use During Periods of HIV Risk Among East African Women in Serodiscordant Relationships | Pyra | 2018 | 10.1097/QAI.0000000000001561 | Prospective Cohort | Serodiscordant Couple | Multiple Countries | Multiple Countries |
| 209 | Fertility Intentions, Pregnancy, and Use of PrEP and ART for Safer Conception Among East African HIV Serodiscordant Couples | Heffron | 2018 | 10.1007/s10461-017-1902-7 | Prospective Cohort | Serodiscordant Couple | Multiple Countries | Multiple Countries |
| 210 | An Empiric Risk Score to Guide PrEP Targeting Among MSM in Coastal Kenya | Wahome | 2018 | 10.1007/s10461-018-2141-2 | Prospective Cohort | MSM | Kenya | Eastern |
| 211 | Patterns of Gender-Based Violence and Associations with Mental Health and HIV Risk Behavior Among Female Sex Workers in Mombasa, Kenya: A Latent Class Analysis | Roberts | 2018 | 10.1007/s10461-018-2107-4 | Prospective Cohort | Sex Worker | Kenya | Eastern |
| 212 | Zimbabwe Population-Based HIV Impact Assessment | ZIMPHIA 2016 | 2017 | NA | Cross Sectional Incidence | General Population | Zimbabwe | Southern |
| 213 | A comparison of four condom-use measures in predicting pregnancy, cervical STI and HIV incidence among Zimbabwean women | Minnis | 2010 | 10.1136/sti.2009.036731 | Prospective Cohort | General Population | Zimbabwe | Southern |
| 214 | Increased Risk of HIV Acquisition among Kenyan Men with Human Papillomavirus Infection | Smith | 2010 | 10.1086/652408 | Secondary Trial Analysis | General Population | Kenya | Eastern |
| 215 | Baseline Factors Associated With Incident HIV and STI in Four Microbicide Trials | Feldblum | 2010 | NA | Clinical Trial | High Risk | Benin, Ghana, Nigeria, South Africa, Uganda | Eastern/Southern/Western/Central |
| 216 | Determinants of Differential HIV Incidence Among Women in Three Southern African Locations | Mavedzenge | 2011 | 10.1097/QAI.0b013e3182254038 | Secondary Trial Analysis | General Population | South Africa, Zimbabwe | Multiple Countries |
| 217 | Increased risk of HIV-1 transmission in pregnancy: a prospective study among African HIV-1-serodiscordant couples | Mugo | 2011 | 10.1097/QAD.0b013e32834a9338 | Prospective Cohort | Serodiscordant Couple | Multiple Countries | Multiple Countries |
| 218 | High-Risk Human Papillomavirus Is Associated with HIV Acquisition among South African Female Sex Workers | Auvert | 2011 | 10.1155/2011/692012 | Secondary Trial Analysis | Sex Worker | South Africa | Southern |
| 219 | Factors Associated With Herpes Simplex Virus Type 2 Incidence in a Cohort of Human Immunodeficiency Virus Type 1-Seronegative Kenyan Men and Women Reporting High-Risk Sexual Behavior | Okuku | 2011 | 10.1097/OQL.0b013e31821a6225 | Prospective Cohort | High Risk | Kenya | Eastern |
| 220 | Disparate Associations of HLA Class I Markers with HIV-1 Acquisition and Control of Viremia in an African Population | Song | 2011 | 10.1371/journal.pone.0023469 | Prospective Cohort | Serodiscordant Couple | Zambia | Southern |
| 221 | Linking HIV prevention and care for community interventions among high-risk women in Burkina Faso--the ARNS 1222 "Yerelon" cohort | Konate | 2011 | 10.1097/QAI.0b013e3182207a3f | Prospective Cohort | Multiple Risk Groups | Burkina Faso | Western/Central |
| 222 | Use of hormonal contraceptives and risk of HIV-1 transmission: a prospective cohort study | Heffron | 2012 | 10.1016/S1473-3099(11)70247-X | Prospective Cohort | Serodiscordant Couple | Multiple Countries | Multiple Countries |
| 223 | Outside Sexual Partnerships and Risk of HIV Acquisition for HIV Uninfected Partners in African HIV Serodiscordant Partnerships | Ndase | 2012 | 10.1097/QAI.0b013e318237b864 | Secondary Trial Analysis | Serodiscordant Couple | Multiple Countries | Multiple Countries |
| 224 | Determinants of Per-Coital-Act HIV-1 Infectivity Among African HIV-1–Serodiscordant Couples | Hughes | 2012 | 10.1093/infdis/jir747 | Secondary Trial Analysis | Serodiscordant Couple | Multiple Countries | Multiple Countries |
| 225 | Assessing and evaluating the combined impact of behavioural and biological risk factors for HIV seroconversion in a cohort of South African women | Wand | 2012 | 10.1080/09540121.2012.687820 | Secondary Trial Analysis | General Population | South Africa | Southern |
| 226 | HLA class I associations with rates of HIV-1 seroconversion and disease progression in the Pumwani Sex Worker Cohort | Peterson | 2013 | 10.1111/tan.12051 | Prospective Cohort | Sex Worker | Kenya | Eastern |
| 227 | Challenges of Diagnosing Acute HIV-1 Subtype C Infection in African Women: Performance of a Clinical Algorithm and the Need for Point-of-Care Nucleic-Acid Based Testing | Mlisana | 2013 | 10.1371/journal.pone.0062928 | Prospective Cohort | High Risk | South Africa | Southern |
| 228 | Effects of hormonal contraceptive use on HIV acquisition and transmission among HIV-discordant couples | Lutalo | 2013 | 10.1097/QAD.0000000000000045 | Prospective Cohort | Serodiscordant Couple | Uganda | Eastern |
| 229 | Impact of an Adherence Intervention on the Effectiveness of Tenofovir Gel in the CAPRISA 004 Trial | Mansoor | 2014 | 10.1007/s10461-014-0752-9 | Secondary Trial Analysis | High Risk | South Africa | Southern |
| 230 | Human Immunodeficiency Virus Seroconversion and Associated Risk Factors among Pregnant Women Delivering at Bugando Medical Center in Mwanza, Tanzania | Mbena | 2014 | 10.4103/2141-9248.141539 | Retrospective Cohort | Pregnant Women | Tanzania | Eastern |
| 231 | The impact of hormonal contraception and pregnancy on sexually transmitted infections and on cervicovaginal microbiota in african sex workers | Borgdorff | 2015 | 10.1097/OLQ.0000000000000245 | Prospective Cohort | Sex Worker | Rwanda | Eastern |
| 232 | Risk Factors for HIV Acquisition in a Prospective Nairobi-Based Female Sex Worker Cohort | McKinnon | 2015 | 10.1007/s10461-015-1118-7 | Prospective Cohort | Sex Worker | Kenya | Eastern |
| 233 | HIV pre-exposure prophylaxis in transgender women: a subgroup analysis of the iPrEx trial | Deutsch | 2015 | 10.1016/S2352-3018(15)00206-4 | Secondary Trial Analysis | Multiple Risk Groups | Multiple Countries | Multiple Countries |
| 234 | Oral and injectable contraceptive use and HIV acquisition risk among women in four African countries: a secondary analysis of data from a microbicide trial | Balkus | 2016 | 10.1016/j.contraception.2015.10.010 | Secondary Trial Analysis | General Population | South Africa, Malawi, Zambia, Zimbabwe | Multiple Countries |
| 235 | Viral and Host Characteristics of Recent and Established HIV-1 Infections in Kisumu based on a Multiassay Approach | Otecko | 2016 | 10.1038/srep37964 | Cross Sectional Incidence | General Population | Kenya | Eastern |
| 236 | Risk of heterosexual HIV transmission attributable to sexually transmitted infections and non-specific genital inflammation in Zambian discordant couples, 1994–2012 | Wall | 2017 | 10.1093/ije/dyx045 | Prospective Cohort | Serodiscordant Couple | Zambia | Southern |
| 237 | Daily and non-daily pre-exposure prophylaxis in African women (HPTN 067/ADAPT Cape Town Trial): a randomised, open-label, phase 2 trial | Bekker | 2018 | 10.1016/S2352-3018(17)30156-X | Clinical Trial | High Risk | South Africa | Southern |
| 238 | Pre-exposure prophylaxis for HIV-negative persons with partners living with HIV: uptake, use, and effectiveness in an open-label demonstration project in East Africa | Heffron | 2017 | 10.12688/gatesopenres.12752 | Secondary Trial Analysis | Serodiscordant Couple | Multiple Countries | Multiple Countries |
| 239 | High Acceptability and Increased HIV-Testing Frequency After Introduction of HIV Self-Testing and Network Distribution Among South African MSM | Lippman | 2018 | 10.1097/QAI.0000000000001601 | Prospective Cohort | MSM | South Africa | Southern |
| 240 | Short Communication: Assessing Estimates of HIV Incidence with a Recent Infection Testing Algorithm That Includes Viral Load Testing and Exposure to Antiretroviral Therapy | Kim | 2018 | 10.1089/AID.2017.0316 | Cross Sectional Incidence | General Population | Kenya, South Africa | Eastern/Southern |
| 241 | Targeted Pregnancy and Human Immunodeficiency Virus Prevention Risk-Reduction Counseling for Young Women: Lessons Learned from Biomedical Prevention Trials | Ramjee | 2018 | 10.1093/infdis/jiy388 | Secondary Trial Analysis | Multiple Risk Groups | South Africa | Southern |
| 242 | The Tablets, Ring, Injections as Options (TRIO) study: what young African women chose and used for future HIV and pregnancy prevention | van der Straten | 2018 | 10.1002/jia2.25094 | Clinical Trial | General Population | Kenya, South Africa | Multiple Countries |
| 243 | Twenty-Year Evolution of Hepatitis B Virus and Human Immunodeficiency Virus Prevalence and Incidence in Voluntary Blood Donors in Côte d’Ivoire | Seri | 2018 | 10.1093/ofid/ofy060 | Prospective Cohort | General Population | Cote d'Ivoire | Western/Central |
| 244 | Transactional sex and incident HIV infection in a cohort of young women from rural South Africa | Kilburn | 2018 | 10.1097/QAD.0000000000001866 | Secondary Trial Analysis | General Population | South | Southern |
| 245 | Estimating Incidence Of HIV Among Adults Visiting A Voluntary Counseling And Testing Centers At Khartoum State, Sudan | Mohammed | 2014 | NA | Cross Sectional Incidence | General Population | Sudan | Eastern |
| 246 | Adult male circumcision as an intervention against HIV: An operational study of uptake in a South African community (ANRS 12126) | Lissouba | 2011 | 10.1186/1471-2334-11-253 | Cross Sectional Incidence | General Population | South Africa | Southern |
| 247 | Identifying Risk Factors for Recent HIV Infection in Kenya Using a Recent Infection Testing Algorithm: Results from a Nationally Representative Population-Based Survey | Kim | 2016 | 10.1371/journal.pone.0155498 | Cross Sectional Incidence | General Population | Kenya | Eastern |
| 248 | Evaluating the BED Capture Enzyme Immunoassay to Estimate HIV Incidence Among Adults in Three Countries in Sub-Saharan Africa | Kim | 2010 | 10.1089/aid.2009.0218 | Cross Sectional Incidence | Multiple Risk Groups | Cote d'Ivoire, Kenya, South Africa | Eastern/Southern/Western/Central |
| 249 | Prevalence of Transmitted HIV Drug Resistance in Botswana: Lessons Learned from the HIVDR-Threshold Survey Conducted Among Women Presenting for Routine Antenatal Care as Part of the 2007 National Sentinel Survey | Bussmann | 2011 | 10.1089/aid.2009.0299 | Cross Sectional Incidence | Pregnant Women | Botswana | Southern |
| 250 | Universal Testing, Expanded Treatment, and Incidence of HIV Infection in Botswana | Makhema | 2019 | 10.1056/NEJMoa1812281 | Clinical Trial | General Population | Botswana | Southern |
| 251 | HIV Testing and Treatment with the Use of a Community Health Approach in Rural Africa | Havlir | 2019 | 10.1056/NEJMoa1809866 | Clinical Trial | General Population | Uganda, Kenya | Multiple Countries |
| 252 | Effect of Universal Testing and Treatment on HIV Incidence — HPTN 071 (PopART) | Hayes | 2019 | 10.1056/NEJMoa1814556 | Clinical Trial | General Population | Multiple Countries | Multiple Countries |
| 253 | HIV incidence among women using intramuscular depot medroxyprogesterone acetate, a copper intrauterine device, or a levonorgestrel implant for contraception: a randomised, multicentre, open-label trial | ECHO Trial Consortium | 2019 | 10.1016/S0140-6736(19)31288-7 | Clinical Trial | General Population | Multiple Countries | Multiple Countries |
| 254 | Effect of ART scale-up and female migration intensity on risk of HIV acquisition: results from a population-based cohort in KwaZulu-Natal, South Africa | Dzomba | 2019 | 10.1186/s12889-019-6494-x | Prospective Cohort | General Population | South Africa | Southern |
| 255 | Time to change the paradigm: limited condom and lubricant use among Nigerian men who have sex with men and transgender women despite availability and counseling | Crowell | 2019 | 10.1016/j.annepidem.2018.12.004 | Prospective Cohort | Multiple Risk Groups | Nigeria | Western/Central |
| 256 | HIV-seroconversion among HIV-1 serodiscordant married couples in Tanzania: a cohort study | Colombe | 2019 | 10.1186/s12879-019-4151-8 | Prospective Cohort | Serodiscordant Couple | Tanzania | Eastern |
| 257 | HIV Retesting of HIV-Negative Pregnant Women in the Context of Prevention of Mother-to-Child Transmission of HIV in Primary Health Centers in Rural Zambia: What Did We Learn? | Mandala | 2019 | 10.1177/2325958218823530 | Prospective Cohort | Pregnant Women | Zambia | Southern |
| 258 | A Window Into the HIV Epidemic from a South African Emergency Department | Hansoti | 2019 | 10.1089/AID.2018.0127 | Cross Sectional Incidence | Other | South Africa | Southern |
| 259 | Tenofovir 1% vaginal gel for prevention of HIV-1 infection in women in South Africa (FACTS-001): a phase 3, randomised, double-blind, placebo-controlled trial | Delany-Moretlwe | 2018 | 10.1016/S1473-3099(18)30428-6 | Clinical Trial | General Population | South Africa | Southern |
| 260 | Persistently high incidence of HIV and poor service uptake in adolescent girls and young women in rural KwaZulu-Natal, South Africa prior to DREAMS | Chimbindi | 2018 | 10.1371/journal.pone.0203193 | Prospective Cohort | General Population | South Africa | Southern |
| 261 | Human Immunodeficiency Virus Incidence Among Women at High-Risk of Human Immunodeficiency Virus Infection Attending a Dedicated Clinic in Kampala, Uganda: 2008-2017 | Kasamba | 2019 | 10.1097/OLQ.0000000000000978 | Prospective Cohort | High Risk | Uganda | Eastern |
| 262 | Age-disparate partnerships and incident HIV infection in adolescent girls and young women in rural South Africa | Stoner | 2019 | 10.1097/QAD.0000000000002037 | Secondary Trial Analysis | General Population | South Africa | Southern |
| 263 | HIV incidence, pregnancy, and implementation outcomes from the Sakh'umndeni safer conception project in South Africa: a prospective cohort study | Schwartz | 2019 | 10.1016/S2352-3018(19)30144-4 | Prospective Cohort | Serodiscordant Couple | South Africa | Southern |
| 264 | Does bacterial vaginosis modify the effect of hormonal contraception on HIV seroconversion | Sabo | 2019 | 10.1097/QAD.0000000000002167 | Prospective Cohort | Sex Worker | Kenya | Eastern |
| 265 | A decade of sustained geographic spread of HIV infections among women in Durban, South Africa | Ramjee | 2019 | 10.1186/s12879-019-4080-6 | Secondary Trial Analysis | General Population | South Africa | Southern |
| 266 | Risk of HIV-1 acquisition among South African women using a variety of contraceptive methods in a prospective study | Palanee-Phillips | 2019 | 10.1097/QAD.0000000000002260 | Secondary Trial Analysis | General Population | South Africa | Southern |
| 267 | Individual and Sexual Network Predictors of HIV Incidence Among Men Who Have Sex With Men in Nigeria | Nowak | 2019 | 10.1097/QAI.0000000000001934 | Prospective Cohort | MSM | Nigeria | Western/Central |
| 268 | Sexual Partner Types and Incident HIV Infection Among Rural South African Adolescent Girls and Young Women Enrolled in HPTN 068: A Latent Class Analysis | Nguyen | 2019 | 10.1097/QAI.0000000000002096 | Secondary Trial Analysis | General Population | South Africa | Southern |
| 269 | Cross-sectional estimates revealed high HIV incidence in Botswana rural communities in the era of successful ART scale-up in 2013-2015 | Moyo | 2018 | 10.1371/journal.pone.0204840 | Cross Sectional Incidence | General Population | Botswana | Southern |
| 270 | Early antiretroviral therapy and daily pre‐exposure prophylaxis for HIV prevention among female sex workers in Cotonou, Benin: a prospective observational demonstration study | Mboup | 2018 | 10.1002/jia2.25208 | Prospective Cohort | Sex Worker | Benin | Western/Central |
| 271 | Village community mobilization is associated with reduced HIV incidence in young South African women participating in the HPTN 068 study cohort | Lippman | 2018 | 10.1002/jia2.25182 | Secondary Trial Analysis | General Population | South Africa | Southern |
| 272 | HIV incidence and predictors of inconsistent condom use among adult men enrolled into an HIV vaccine preparedness study, Rustenburg, South Africa | Maenetje | 2019 | 10.1371/journal.pone.0214786 | Prospective Cohort | Multiple Risk Groups | South Africa | Southern |
| 273 | HIV incidence among pregnant and postpartum women in a high prevalence setting | Machekano | 2018 | 10.1371/journal.pone.0209782 | Prospective Cohort | Pregnant Women | Lesotho | Southern |
| 274 | HIV incidence during breastfeeding and mother-to-child transmission in Cape Town, South Africa | le Roux | 2019 | 10.1097/QAD.0000000000002224 | Prospective Cohort | Pregnant Women | South Africa | Southern |
| 275 | PrEP interest and HIV‐1 incidence among MSM and transgender women in coastal Kenya | Kimani | 2019 | 10.1002/jia2.25323 | Prospective Cohort | Multiple Risk Groups | Kenya | Eastern |
| 276 | Primary HIV prevention in pregnant and lactating Ugandan women: A randomized trial | Homsy | 2019 | 10.1371/journal.pone.0212119 | Clinical Trial | Multiple Risk Groups | Uganda | Eastern |
| 277 | Implementation of a comprehensive safer conception intervention for HIV‐serodiscordant couples in Kenya: uptake, use and effectiveness | Heffron | 2019 | 10.1002/jia2.25261 | Prospective Cohort | Serodiscordant Couple | Kenya | Eastern |
| 278 | Population-level HIV incidence estimates using a combination of synthetic cohort and recency biomarker approaches in KwaZulu-Natal, South Africa | Grebe | 2018 | 10.1371/journal.pone.0203638 | Cross Sectional Incidence | General Population | South Africa | Southern |
| 279 | High HIV incidence and low uptake of HIV prevention services: The context of risk for young male adults prior to DREAMS in rural KwaZulu-Natal, South Africa | Baisley | 2018 | 10.1371/journal.pone.0208689 | Prospective Cohort | General Population | South Africa | Southern |
| 280 | Tsogolo la Thanzi: A Longitudinal Study of Young Adults Living in Malawi's HIV Epidemic | Yeatman | 2019 | 10.1111/sifp.12080 | Prospective Cohort | General Population | Malawi | Southern |
| 281 | Simulated vaccine efficacy trials to estimate HIV incidence for actual vaccine clinical trials in key populations in Uganda | Abaasa | 2019 | 10.1016/j.vaccine.2019.02.072 | Prospective Cohort | Multiple Risk Groups | Uganda | Eastern |
| 282 | Impact of combination HIV interventions on HIV incidence in hyperendemic fishing communities in Uganda: a prospective cohort study | Kagaayi | 2019 | 10.1016/S2352-3018(19)30190-0 | Prospective Cohort | Fisher Folk | Uganda | Eastern |
| 283 | Malawai Population-Based HIV Impact Assessment | MPHIA 2016 | 2018 | NA | Cross Sectional Incidence | General Population | Malawi | Southern |
| 284 | Zambia Population-based HIV Impact Assessment | ZAMPHIA 2016 | 2018 | NA | Cross Sectional Incidence | General Population | Zambia | Southern |
| 285 | Cameroon Population-based HIV Impact Assessment | CAMPHIA 2017 | 2018 | NA | Cross Sectional Incidence | General Population | Cameroon | Western/Central |
| 286 | Ethopia Population-based HIV Impact Assessment | EPHIA 2017-2018 | 2018 | NA | Cross Sectional Incidence | General Population | Ethopia | Eastern |
| 287 | Swaziland HIV Incidence Measurement Survey 2 (SHIMS2): a prospective national cohort study | SHIMS2 2016-2017 | 2018 | NA | Cross Sectional Incidence | General Population | Ethopia | Southern |
| 288 | Lesotho Population-based HIV Impact Assessment | LePHIA 2016-2017 | 2018 | NA | Cross Sectional Incidence | General Population | eSwatini | Southern |
| 289 | Namibia Population-based HIV Impact Assessment | NAMPHIA 2017 | 2018 | NA | Cross Sectional Incidence | General Population | Lesotho | Southern |
| 290 | Tanzania HIV Impact Survey | THIS 2016-2017 | 2018 | NA | Cross Sectional Incidence | General Population | Tanzania | Eastern |
| 291 | Uganda Population-based HIV Impact Assessment | UPHIA 2016-2017 | 2019 | NA | Cross Sectional Incidence | General Population | Uganda | Eastern |
| 292 | South Africa National HIV Prevalence, Incidence, Behavior, and Communication Survey | SABSSM | 2018 | NA | Cross Sectional Incidence | Multiple Risk Groups | South Africa | Southern |
| *Veldhuijzen 2010 - 96% of participants were SW, Vanderpitte 2013 - 95% of participants were SW | | | |  |  |  |  |  |
| Other includes defined groups of people (i.e., farm/plantation workers, police officers, patients with TB, patients in ED etc). | | | | | |  |  |  |
